# A Virtual Patients Ensemble Approach for Predicting Surgical Complications

**DOI:** 10.1101/2025.09.21.25336262

**Authors:** Yair Neuman, Yochai Cohen, Yiftach Neuman

## Abstract

AI has shown promise in predicting surgical complications, but most existing models estimate overall risk levels rather than identifying the specific complications an individual patient may develop. We present an AI agent that uses a *Virtual Patients Ensemble (VPE)* approach to generate individualized predictions of surgical complications from unstructured case descriptions. The agent applies structured reasoning to extract diagnoses, surgical procedures, and risk factors from clinical narratives. From this profile, it generates a *cohort of N virtual patients*, each a plausible variation of the original case. This ensemble captures uncertainty in patient-specific risk factors and grounds LLM-based clinical reasoning in individualized clinical scenarios. For each virtual patient, the agent predicts the most likely complications, and a final distribution is presented over the virtual patients. The agent was evaluated on 1440 case reports from the PMC-Patients dataset, of which 186 met the inclusion criteria. Predictive performance was compared with both null-hypothesis expectations and baseline LLM predictions. The agent correctly identified 32% of the observed complications, significantly outperforming the null-hypothesis baseline and a baseline prediction generated by the LLM. Unlike risk calculators or machine-learning models trained on population averages, this approach derives predictions directly from a patient’s clinical profile, generating a VPE to predict specific complications rather than general risk levels. The results suggest that ensemble-based, patient-centered simulation can support clinical decision-making by offering interpretable, individualized predictions. Prospective validation is required before integration into practice. We thus provide surgeons with an app for experimenting with the agent and providing feedback for improvement.

## Introduction

Surgeons face significant challenges when anticipating postoperative complications. A patient’s clinical profile often includes a complex interplay of medical history, comorbidities, and procedure-related risks, making individualized prediction difficult. Traditional approaches, such as the ACS Surgical Risk Calculator^1^, have demonstrated value compared with physician intuition^2^, yet these models that focus on risk are constrained by linear assumptions in estimating log odds and limited capacity to capture complex interactions.

In recent years, artificial intelligence (AI) has emerged as a promising tool for surgical risk prediction^3^. Most efforts have focused on machine-learning models trained on population-level datasets^4^. While these models achieve reasonable accuracy, their reliance on population averages can limit their ability to provide patient-specific predictions. Systematic reviews of AI for predicting postoperative complications after major surgery highlight both potential and variability in performance^5,6,7^, with recent work noting “variable rates of precision”^8^ and “inconsistent results”^9^.

More recently, large language models (LLMs) have shown potential for medical applications^10,11^ because of their ability to perform context-sensitive^12^ and probabilistic reasoning^13^. A few studies demonstrate their capacity to classify complications or apply clinical grading systems. For example^14^, showed that ChatGPT-4 can grade postoperative complications using the Clavien–Dindo classification with promising accuracy. Yet applications in surgery remain at an early stage^15,16^.

We present an AI agent for predicting surgical complications. Instead of estimating general risk levels or restricting predictions to a *predefined* and *closed* set of complications, the agent generates an *open-ended* distribution of likely complications. This process involves the generation of a Virtual Patients Ensemble (VPE): a cohort of simulated patient profiles derived from a single clinical case. By varying risk factors across simulated patients, the AI agent explores plausible outcome scenarios and identifies complications most likely to occur. Therefore, the AI agent combines the clinical breadth of LLM reasoning with individualized patient simulation, offering interpretable predictions that align with precision medicine.

### The AI agent

The agent, called BIBAS, accepts the patient’s description as input. Prompt 1 simulates a panel of experts to identify a diagnosis leading to a surgical procedure that produces an intra- or postoperative complication. Prompt 1.5 identifies the MeSH Tree Number of the complication. Prompt 2 identifies the text preceding the surgical procedure, and Prompt 3 identifies in this text those of the patient’s risk factors that may increase the likelihood of a surgical complication. Prompts 4A-4C simulate the VPE. The simulation uses the patient’s specific profile, which includes diagnosis, surgical procedure, risk factors, and expected timing of a complication. It clones up to 50 virtual patients from this profile. We generated 50 virtual patients as a pragmatic choice, balancing computational efficiency with performance, but the number of patients is a variable parameter in the model and can be changed by the user when running the simulation. The justification for generating 50 virtual patients appears in the appendix. All virtual patients share the same diagnosis and surgical procedure, but differ in the parameters of the risk factors. For each virtual patient, the system predicts the most likely complications that share the same time frame as the observed complication (i.e., immediate or early complication versus late or long-term). The system then summarizes the distribution of the identified complications across the VPE and presents them in descending order of probability.

### Justifying the Virtual Patients Ensemble approach

The justification for using a VPE is fourfold:

#### 1. Patient-centered modeling rather than population averages

Traditional risk calculators and most ML models are trained on population-level data. While they perform well statistically, they struggle with individualization. A VPE starts from the unique patient profile (diagnosis, surgical plan, and risk factors) and generates multiple patient-specific simulations. This approach aligns more closely with precision medicine by focusing on the *individual* rather than the *average*.

#### 2. Captures complex interactions between risk factors

Surgical outcomes emerge from interactions (e.g., advanced age × infection × comorbidities) that are often oversimplified by linear models and even by some ML approaches. By varying the patient’s risk factor parameters across virtual patients, the VPE explores a wide range of plausible scenarios, enabling the emergence of complication patterns that depend on non-linear interactions.

#### 3. Transparency and clinical interpretability

Surgeons may find it difficult to trust or interpret black-box predictions. A VPE produces a distribution of complications across virtual patients—for example, “40% of the ensemble developed surgical site infection.” This frequency-based output is intuitive for clinicians and mirrors their own reasoning when they mentally simulate “patients like this one” to anticipate complications.

#### 4. Bridging LLM knowledge with clinical data

Large language models (LLMs) bring broad clinical knowledge but lack grounding in structured, patient-specific variability. A VPE bridges this gap: it anchors the LLM’s knowledge and general reasoning in concrete patient features while allowing exploration of uncertainty across multiple plausible scenarios. The result is a prediction framework that is both knowledge-driven and individualized.

### Illustrating the agent’s outcomes

The patient descriptions include the case of Maria. This case provides the following information: “Maria G., a 74-year-old female, presented to the emergency department with a 2-day history of severe epigastric pain radiating to the back, associated with nausea and multiple episodes of non-bloody vomiting.” CT imaging confirmed the diagnosis of “acute gallstone pancreatitis, with signs of peripancreatic fat stranding and inflammation.” The surgical intervention was “laparoscopic cholecystectomy,” followed on postoperative day 2 by the complication, viz., a “postoperative localized intra-abdominal infection”. BIBAS identified the following risk factors:

> [Age 74 years, Demographic]
>
> [Acute pancreatitis, Anatomical/Pathological]
>
> [Elevated white blood cell count, Laboratory/Physiological]
>
> [Peripancreatic inflammation, Anatomical/Pathological]

And through the VPE simulation, the top predicted complications were:

1. Surgical site infection
2. Bile duct injury
3. Postoperative bleeding

### Evaluation

#### Sample

We used the PMC-Patients dataset^17^ and ran the agent on 1440 patient descriptions. The latter were randomly sampled from each of six age groups (see Table 1 in the appendix), with equal weight given to the proportion of males and females. The data set was collected from the case reports in the PMC archive. Case reports naturally emphasize the rare, unusual, or unique^18,19^. Therefore, the use of this dataset is particularly challenging, given that we analyze free texts rather than a structured dataset, and texts dealing with case reports where the distribution of complications may deviate significantly from baseline predictions.

#### Procedure

All prompts appear in the appendix. GPT-5 was the LLM used for all prompts, and we ran the agent through the API. We applied Prompt 1 and identified cases of a diagnosis leading to surgical intervention and subsequent complications. In this study, we analyzed only surgical procedures that led to a complication. Next, we selected cases where the text preceding the surgical procedure (i.e., the context) was longer than 400 characters. This ensured enough contextual information to identify risk factors. 186 patient descriptions were selected for the final analysis. A case report may include more than one surgical intervention and more than one complication resulting from a given intervention. For brevity, we analyzed the first surgical intervention mentioned in the text, followed by a single complication, and focused on the first complication resulting from the surgery.

#### Analysis and Results

We performed three tests to evaluate the performance of BIBAS.

#### Primary Evaluation

We looked for a match between the MeSH tree number of the observed complication and the tree numbers of each of the ten highest-ranked complications predicted by the agent. We focused on ten complications because the patient descriptions may be biased toward rarer complications. We tested whether one of the ten predicted complications was the same as the observed complication (i.e., a HIT). There were 59 cases of HIT (32%) with 95% CI [0.25, 0.39], and 40 of these cases (68%) were in the top three predicted complications. We compared this hit rate to a random guess and a baseline LLM prediction. To compare this hit rate to a random guess, we identified 269 unique complications predicted by the agent. The chance of a match between a randomly selected complication from this list and the observed complication was therefore 0.004. The chance of a match between the observed complication and one of the ten predicted complications was p = 0.04 with 95% CI [0.02, 0.079]. The difference between the proportion of complications identified by BIBAS and the one hypothesized under the null hypothesis was found to be statistically significant (z = 7.03, p < 0.001). The effect size (Cohen’s h = 0.80) was found to exceed Cohen’s convention for a medium effect size (h = 0.5). We used Prompt 5 to identify the most likely complication to follow the surgical procedure (MOST_LIKELY). This predicted complication served as a baseline prediction generated by the LLM. Next, we looked for a match between this LLM prediction and what was observed. There were 24 cases of HIT (13%). This means that the proportion of cases identified by BIBAS is about 2.5 times greater than the proportion identified by the LLM baseline prediction. The difference between the hit rate of BIBAS and the baseline prediction generated by the LLM was statistically significant (z = 4.39, p < 0.001, Cohen’s h = 0.46)

#### Secondary Evaluation 1

We measured how similar the meaning of each predicted complication was to the meaning of the observed complication. To address this challenge, we represented the meaning of a complication by using its text embedding. More specifically, we used OpenAI’s text-embedding-3-large model, which includes 3072 dimensions^20^. Next, we measured the cosine similarity between each of the ten highest predicted complications and the observed complications and selected the predicted complication that scored highest (HIGHEST_PREDICTED). This prediction is BIBAS’s best guess in terms of semantic similarity. Next, we measured the cosine similarity between the embedding of MOST_LIKELY and the observed complication. We hypothesized that, if BIBAS performs better than a general prediction generated by the LLM, then its best prediction, in terms of semantic similarity to the observed (i.e., HIGHEST_PREDICTED), should be closer to the meaning of the observed complication than the meaning of the MOST_LIKELY. The average HIGHEST_PREDICTED score was 0.49(0.12), with 95% CI [0.48–0.51], and for the MOST_LIKELY it was 0.41 (0.11) with a 95% CI [0.39–0.42]. We computed the difference between the two scores, expecting it to be significantly higher than 0. To test this hypothesis, we used the one-sample Wilcoxon signed-rank test. In 75% of the cases, the difference in the similarity score was positive and in favor of HIGHEST_PREDICTED. The test was found to be statistically significant (V = 13830, p < 0.001) and with a medium-large effect size (rank-biserial correlation = 0.59).

#### Secondary Evaluation 2

Finally, through Prompt 6, we tested whether the HIGHEST_PREDICTED and the observed complication share the same complication category. We used 13 categories of complications. Running Prompt 6, we found a match in 138 out of 186 cases (74%). Given the expected proportion of complications sharing the same category under the null hypothesis (i.e., 0.08), the difference was found to be statistically significant (z = 34.016, p < 0.001) and with a very large effect size (Cohen’s h = 1.51). When we compared the proportion of surgical categories shared by MOST_LIKELY and the observed (i.e., 36%) to the shared categories with HIGEST_PREDICTED (i.e., 74%), the difference was found to be statistically significant (z = 7.36, p < 0.001, Cohen’s h = 0.78).

## Conclusions

This study introduces a novel framework for predicting surgical complications using a VPE. To our knowledge, this is the first approach that combines large language model reasoning with an ensemble of patient-specific simulations to move beyond general +risk estimation and predict *concrete complications*. Unlike existing methods that rely on population-level averages or static ML models, the VPE framework bootstraps *directly* from a patient’s clinical description, generates a cohort of individualized virtual patients, and identifies complication patterns emerging from complex interactions of risk factors. The significance of this contribution lies in its dual novelty: (1) reframing surgical complication prediction as an individualized, scenario-based simulation problem rather than a population-based probability estimate, and (2) operationalizing LLM knowledge in a structured, clinically transparent way. Nevertheless, the agent requires human validation, which was beyond the budget scope of the current paper, and further validation in prospective and structured clinical datasets is required. Moreover, running the simulation is computationally expensive, so we provide a “lightweight” prompt in the appendix that can be run directly from the UI. This version performs well, although it does not generate the VPE, but it works *as if* it does (see the appendix). We also provide a link to an app running the agent from the API^21^. In sum, the agent and the approach have the potential to augment surgical decision-making and broaden the toolkit of predictive methods available to clinicians. In its present state, the agent can be used by surgeons as an assisting tool for mapping likely complications.

## Data Availability

All data produced in the present work are contained in the manuscript

https://doi.org/10.6084/m9.figshare.30075007

## Data sharing

The dataset generated and analyzed for this study is freely available at: https://doi.org/10.6084/m9.figshare.30075007

## Acknowledgments

The authors would like to thank Prof. Irun Cohen, MD, for his helpful feedback and Dr. Dan Assaf, MD, for constructive talks.

## Authors’ contribution

Y.N. proposed the original approach.

Y.N., Y.C. Yi. N. developed the methods.

Y.C. Yi. N. wrote the code for the analysis.

Y.N., Y.C., Yi. N. wrote the manuscript.

Yi. N. designed the app.

