## supplementary appendix for "A Virtual Patients Ensemble Approach for Predicting Surgical Complications"

Table of Contents

Determining the number of Virtual Patients

BIBAS prompts:

Prompt 1

Prompt 1.5

Prompt 2

Prompt 3

Prompts 4A, 4B and 4C

Table 1. Age Groups

Prompt 5

Prompt 6

BIBAS UI version

**Determining the number of Virtual Patients**

First, it is important that we present the 10 most likely complications, and therefore our focus is on ranking which is more robust to sampling variability. For each real patient in the cohort (n = 186), we generated 50 virtual patients. The choice of 50 virtual balances computational efficiency and statistical considerations. The number of virtual patients per patient was determined by the hypothesized prevalence of the rarest complication (i.e., 2%), ensuring each real patient generated an expected value of 1.0 events for rare complications (50 × 0.02 = 1.0). This approach produced 9,300 total virtual patients across the cohort, yielding approximately 186 expected events for the rarest complication. Therefore, for complication ranking, 50 virtual patients per real patient provides an optimal balance between discriminative resolution and computational efficiency. This sample size generates sufficient frequency resolution to reliably rank-order the 10 complications while maintaining stability in relative risk assessment. Even for rare complications (2% prevalence), the expected 1 event per patient provides an adequate signal to distinguish it from both higher-risk complications (3-7 events) and lower-risk complications (0 events) in the ranking hierarchy.

**Prompt 1.**

**Expert Panel Simulation to Identify Diagnoses, Surgeries, and Complications in a Patient’s Clinical Description**

**Definitions:**

1. **Surgical Diagnosis** is a diagnosis that:
   - Has a **direct causal relationship** to a **surgical intervention**.
   - Is **documented in a clinical description** as the **main reason** for performing the surgical intervention.
2. **Surgical Intervention** is a procedure that:
   - Involves **incision, puncture, catheterization, or internal access** to diagnose, treat, or correct a medical condition.
   - Includes **therapeutic and diagnostic procedures** such as open surgeries, laparoscopic/endoscopic procedures, interventional techniques (e.g., stent placement, coronary angiography, biopsies), emergency decompressions, or diagnostic biopsies.
   - **Note**: Diagnostic catheterizations and angiographies involving vascular access and internal instrumentation qualify as surgical interventions.
3. **Surgical Complication** is any adverse medical outcome that:
   - Occurs **intraoperatively or postoperatively (within or after 30 days)**,
   - Is **causally or temporally associated** with a **surgical intervention**,
   - Represents an **undesirable effect, consequence, or failure of recovery**.

✅ **Examples include:**

- Postoperative infections (e.g., intra-abdominal abscess, fluid collection, wound infection)
- Postprocedural bleeding or bile leak
- Surgical site inflammation
- Digestive complications (e.g., ileus, leaks, obstruction)
- Other defined postprocedural disorders

**The Typology of Complications**

**1. Immediate Complications**

- **Timing:** During surgery or within **24 hours**
- **Examples:**
  - Hemorrhage (intraoperative or postoperative bleeding)
  - Anesthesia-related issues (e.g., aspiration, cardiac events)
  - Injury to adjacent structures
  - Acute allergic reactions

**🕓 2. Early Complications**

- **Timing:** Within **24 hours to 30 days** after surgery
- **Examples:**
  - Surgical site infection (SSI)
  - Pneumonia
  - Deep vein thrombosis (DVT) or pulmonary embolism (PE)
  - Paralytic ileus or bowel obstruction
  - Urinary leakage or fistula
  - Acute kidney injury
  - Wound dehiscence

**🗓️ 3. Late Complications**

- **Timing:** After **30 days**, often up to **months post-op**
- **Examples:**
  - Anastomotic strictures
  - Hernia at the surgical site
  - Ureteral obstruction or stricture
  - Chronic pain
  - Delayed infections (e.g., deep abscess)

**📆 4. Long-term (or Chronic) Complications**

- **Timing:** **Months to years** after surgery
- **Examples:**
  - **Neoplasm recurrence**
  - Chronic kidney disease (from urinary diversion)
  - Metabolic disturbances (e.g., acidosis with ileal conduit)
  - Sexual dysfunction
  - Long-term urinary incontinence or retention

**Objective:**

Simulate a panel of **seven expert surgeons** analyzing a patient’s clinical description. Each expert independently identifies:

- **Surgical Interventions**
- The **Diagnosis** that led to each surgical intervention
- The **Complication**, if any, that followed the surgical intervention

**Expert Task:**

Each expert independently:

1. Reads the patient’s clinical description.
2. Determines whether a **surgical intervention** is explicitly mentioned.
   - **Answer: YES or NO**
3. If **YES**:
   - Identifies the **primary diagnosis** that led to the surgical intervention.
   - Determines if a **complication** occurred.
     - **Answer: YES or NO**
   - Identifies each complication and labels it as “Immediate”, “Early”, “Late”, or “Long term”.
4. For each identified item (diagnosis, surgical intervention, complication), the expert maps the relevant **MeSH Name descriptor/s** using the **current official MeSH hierarchy**.

**MeSH Tree Structure Guidelines:**

- Use **Medical Subject Headings (MeSH)** from the **current version maintained by the National Library of Medicine**.
- MeSH descriptors are organized into 16 hierarchical categories (e.g., A for anatomic terms, C for diseases, E for procedures).
  - **Each descriptor must be verified** to ensure the accuracy of the term
- **Do not infer tree structure** based on assumed hierarchy or naming patterns.
- Use the **most specific available MeSH descriptor** that precisely represents the concept.
- Exclude from the output the following MeSH descriptors:

Intraoperative complications.

Postoperative complications

**Output Template:**

**1. Surgical Intervention Identification**

- Has a Surgical Intervention been identified? **YES / NO**
- If YES, list each surgical intervention identified by a **majority (≥4 of 7 experts)**:
  - **Surgical Intervention Name**
  - **MeSH descriptor**

**2. Main Diagnosis Leading to Surgical Intervention**

For each intervention identified:

- **Surgical Intervention**: [Intervention Name]
  - **Main Diagnosis**: [Causal Diagnosis]
  - **MeSH descriptor(s)**: [MeSH name]

**3. Complication(s)**

- Has a complication been identified? **YES / NO**
- For each complication related to the intervention:
  - **Complication Name**
  - **MeSH descriptor(s)**

**✅ Internal Quality Control:**

Perform an internal quality check to ensure:

- Each diagnosis, surgical intervention, and complication is **explicitly stated** in the clinical text — **no speculation or assumptions**.
- Each element **matches the definitions** of diagnosis, intervention, or complication per this prompt.
- Ensure the identified surgical complication is a preoperative or postoperative complication, and NOT an ADVERSE OUTCOME.
- There is a **clear causal or temporal relationship**:
  - Diagnosis ➜ Intervention
  - Intervention ➜ Complication (if any)
- Each element is identified by a **majority of the expert panel (≥4 of 7)**
- You MUST ensure that each **MeSH descriptor is valid**

**Notes:**

- Do not show individual expert opinions or reasoning steps.

**Input: [**The patient’s clinical description HERE]

**Example 1:**

**The patient’s description is:**

Maria G., a 74-year-old female, presented to the emergency department with a 2-day history of severe epigastric pain radiating to the back, associated with nausea and multiple episodes of non-bloody vomiting. She reported decreased appetite and subjective fever. Her past medical history included gallstones, hyperlipidemia, and coronary artery disease. On physical examination, the patient was febrile (38.7°C), with tenderness in the right upper quadrant and epigastrium. Laboratory findings revealed elevated white blood cell count, increased serum lipase (1120 U/L), and mildly abnormal liver function tests. Abdominal ultrasound showed gallstones without ductal dilatation, while CT imaging confirmed acute gallstone pancreatitis, with signs of peripancreatic fat stranding and inflammation. The patient was admitted and initially managed with intravenous fluids, analgesia, and bowel rest. On hospital day 3, due to persistent symptoms and recurrent pain, the surgical team proceeded with a laparoscopic cholecystectomy. Intraoperative findings included an inflamed gallbladder with multiple small stones. The surgery was uneventful. However, on postoperative day 2, the patient developed fever (39.1°C), tachycardia, and mild hypotension. Repeat labs showed worsening inflammatory markers. A CT scan of the abdomen revealed a small fluid collection in the gallbladder fossa, consistent with a postoperative localized intra-abdominal infection. The patient was managed conservatively with intravenous antibiotics (piperacillin-tazobactam); no drainage procedure was required. She gradually improved and was discharged on postoperative day 7.

**Output:**

**1. Surgical Intervention Identification**

- **Has a Surgical Intervention been identified? YES**
- **Identified Surgical Intervention(s) (majority ≥4/7):**
  - **Laparoscopic Cholecystectomy — *Cholecystectomy, Laparoscopic***

**2. Main Diagnosis Leading to Surgical Interventions**

- **Laparoscopic Cholecystectomy**
  - **Main Diagnosis: Gallstone Pancreatitis**
  - **MeSH descriptor(s):**
    - ***Cholelithiasis***
    - ***Pancreatitis***

**3. Complication(s)**

- **Laparoscopic Cholecystectomy — YES**
  - **Complication Name: Intra-abdominal infection (localized fluid collection in gallbladder fossa)**
  - **MeSH descriptor(s):**
    - ***Abscess***
    - ***Intra-abdominal Infections***
  - **Timing Classification: Early Complication (postoperative day 2)**

**Example 2.**

**The patient’s description:**

A 38-year-old woman, with a 16-year history of diabetes mellitus, underwent a simultaneous pancreas–kidney transplantation from a blood group and human leucocyte antigen (HLA) A2- and HLA A8-compatible cadaveric donor on 11 September 2001. The procedure was uneventful, and immediate graft function was excellent. The immunosuppressive regimen included timoglobulin (five doses of 1.25 mg/kg/day), tacrolimus (0.2 mg/kg/day in two divided doses), mycophenolate mofetil (2 g/day), and prednisone with a decreasing dosing schedule until 5 mg daily at 6 months. She received 3-month anti-cytomegalovirus (CMV) prophylaxis with oral ganciclovir because of CMV+ donor/CMV− recipient, as well as cotrimoxazole and itraconazole for 6 months according to the protocol of our hospital, and low-molecular-weight heparin during the first 2 weeks followed by acetylsalicylic acid 150 mg/day.\nThe patient was discharged from the hospital on Day 23 after transplantation, with a serum creatinine concentration of 1.2 mg/dL, blood glucose of 83 mg/dL, amylase of 50 mg/dL, and lipase of 35 mg/dL. One month after transplantation, the patient presented with hyperglycaemia that required insulin therapy. Pancreatic sonography and an ultrasound-guided biopsy of the pancreas were unrevealing; tacrolimus was substituted with sirolimus. Blood glucose levels were normal. Five months after transplantation, she had an episode of obstructive uropathy secondary to distal ureteral stenosis, which resolved with balloon dilatation followed by placement of a Double-J catheter. Four years after transplantation, the patient was admitted to the hospital because of renal function deterioration. Renal echography was normal, and plasma levels of sirolimus were within the therapeutic range. Renal biopsy revealed a tubulointerstitial lymphocytic inflammatory infiltrate in 25% of the sample, with more than four lymphocytes per section and cytomegalia. BKV DNA (170,000 copies/mL) was detected by real-time PCR, and immunohistochemical staining for SV-40 was positive. Mycophenolate mofetil was discontinued, and treatment with four doses of cidofovir was instituted. BK viraemia persisted, and 4 months later, haemodialysis was required due to the decline in renal function. In the presence of intermittent BK viraemia and to perform a second kidney transplant, the renal graft was removed in May 2008. The surgical specimen showed the lymphocytic inflammatory infiltrate with SV-40 immunohistochemical positivity. An abdominal computer tomography (CT) scan disclosed bladder wall thickening. Cystoscopy demonstrated a large mass; the biopsy of which revealed adenocarcinoma. Immunohistochemical staining for SV-40 was intensely positive in neoplastic cells but negative in non-neoplastic cells. Radical cystectomy with hysterectomy and bilateral salpingo-oophorectomy was performed. Immunochemical staining of the surgical specimen showed positivity for PI3KC, phospho-AKT, p mTOR, p70S6K, pp70S6K, TGFβ, VEGF, and p53, and was negative for PTNE expression. Post-operatively, PCR BKV DNA assays were repeatedly negative. At follow-up, 1 year after cystectomy, the patient remained on dialysis, with normal pancreatic function, with negative BK viraemias, and with no evidence of metastasis. At 18 months after cystectomy, the patient was admitted because of a pelvic mass in the renal post-transplantation territory, which was successfully removed. Histopathological examination revealed a tumour recurrence with positive immunohistochemical staining for SV-40.

**OUTPUT:**

**Final Output**

**1. Surgical Intervention Identification**

- **Has a Surgical Intervention been identified?** YES
- **Identified Surgical Interventions (majority ≥4/7):**
  - Simultaneous Pancreas–Kidney Transplantation — *Pancreas Transplantation*, *Kidney Transplantation*
  - Balloon Dilatation of Distal Ureteral Stenosis — *Balloon Dilatation*
  - Removal of Renal Graft — *Kidney Transplantation*
  - Radical Cystectomy with Hysterectomy and Bilateral Salpingo‑Oophorectomy — *Cystectomy*, *Hysterectomy*

**2. Main Diagnosis Leading to Surgical Interventions**

- **Simultaneous Pancreas–Kidney Transplantation**
  - **Main Diagnosis:** Diabetes Mellitus (implying kidney failure)
  - **MeSH descriptor(s):** *Diabetes Mellitus*, *Kidney Failure, Chronic*
- **Balloon Dilatation of Distal Ureteral Stenosis**
  - **Main Diagnosis:** Ureteral Obstruction due to distal ureteral stenosis
  - **MeSH descriptor(s):** *Ureteral Obstruction*
- **Removal of Renal Graft**
  - **Main Diagnosis:** Kidney Failure (BK virus-associated nephropathy)
  - **MeSH descriptor(s):** *Kidney Failure, Chronic*, *Polyomavirus Infections*
- **Radical Cystectomy with Hysterectomy and Bilateral Salpingo‑Oophorectomy**
  - **Main Diagnosis:** Urinary Bladder Neoplasm (adenocarcinoma)
  - **MeSH descriptor(s):** *Urinary Bladder Neoplasms*

**3. Complication(s)**

- **Simultaneous Pancreas–Kidney Transplantation** — NO
- **Balloon Dilatation of Distal Ureteral Stenosis** — NO
- **Removal of Renal Graft** — NO
- **Radical Cystectomy with Hysterectomy and Bilateral Salpingo‑Oophorectomy** — YES
  - **Complication Name:** Tumor recurrence (pelvic mass)
  - **MeSH descriptor(s):** *Urinary Bladder Neoplasms* (recurrence)
  - **Timing Classification:** Late Complication

Top of Form

Bottom of Form

**Prompt 1.5**

**Identifying the Specific MeSH Tree Number for a Surgical Complication**

**Task Objective:**

You are given the MeSH descriptors of a surgical intervention and the surgical complications that followed it.

Identify the most specific **MeSH tree number** that best describes each complication.

Return ONLY ONE MeSH tree number that best describes the surgical complication.

**Input:**

1. **Surgical Intervention**: [Surgical Intervention MeSH Descriptor here]
2. **MeSH Complication Descriptor**: [Observed complication MeSH Descriptor here]
3. **Timing Classification of the Complication:** [Timing Classification Here]

**Note: The Timing Classification of the Complication concerns its timing of appearance as follows:**

IMMEDIATE Complications: During surgery or within 24 hours

EARLY Complications: Within 24 hours to 30 days after surgery

LATE Complications: After 30 days, often up to months post-op

LOMG-TERM (or Chronic) Complications: Months to years after surgery

**Process:**

1. **Understand the Relationship Between Surgical Intervention and Complication:**
   - Each surgical intervention may have specific complications associated with it.
   - For each complication, a corresponding MeSH tree number describes it.
   - The **MeSH tree number** hierarchy organizes medical terms, so the more specific the complication, the deeper it is in the hierarchy.
2. **Use the <MeSH Tree Data>**
3. **Determine the Most Specific MeSH Tree Number:**
   - Given a **surgical intervention** and a **MeSH complication descriptor**, the goal is to look up the complication in the **mesh_tree_data** dictionary.
   - Find the complication with the most specific **MeSH tree number**.
   - Return ONLY a single tree number.
   - Use clinical medical knowledge to ensure the MeSH tree number is the most specific and accurate representation of the given surgical complication.
   - Return a dictionary with each **complication descriptor** as the key and its **MeSH tree number** as the value.
4. **IMPORTANT:**

- NEVER return the MeSH descriptors or tree numbers corresponding to the general **descriptors** "postoperative complications" or "intraoperative complications".
- If the most accurate and specific MeSH descriptor and tree number do **not** appear in the **<MeSH Tree Data>,** search it in the **broader MeSH hierarchy** (outside the given subset), specifically in categories **C** (i.e., Diseases) and **E** (i.e., Analytic, Diagnostic, and Therapeutic Techniques)
- The **MeSH tree number** should be chosen based on clinical knowledge and the hierarchy of terms in MeSH.
- If multiple descriptors describe a complication, use the most specific tree number.

5. **Internal quality check:**
   - Perform an internal quality check.
   - Ensure the complication is described using the valid tree number.
6. **Return Format:**
   The output should be the following format: [**Complication descriptor**, **MeSH tree number**
7. **Error Handling:**
   - If a complication is not found under the specified intervention, return a message saying no matching complications were found.

**Example:**

**Input:**

**Surgical Intervention**: [Laparoscopic Cholecystectomy]

**MeSH Complication Descriptor**: [Intra-abdominal infection, Abscess]

**Timing Classification of the Complication: [EARLY]**

**Output:**

[Abdominal Abscess, C01.830.025.020]

\###

**<MeSH Tree Data>**

Postoperative complications MeSH

1. Pathological Conditions, Signs and Symptoms [C23]
2. Pathologic Processes [C23.550]
3. Acantholysis [C23.550.035]
4. Arrhythmias, Cardiac [C23.550.073]
5. Ascites [C23.550.081]
6. Atrial Remodeling [C23.550.113]
7. Azotemia [C23.550.145]
8. Cardiotoxicity [C23.550.161]
9. Channelopathies [C23.550.177]
10. Chromosome Aberrations [C23.550.210]
11. Death [C23.550.260]
12. Dehydration [C23.550.274]
13. Delayed Graft Function [C23.550.277]
14. Disease [C23.550.288]
15. Disease Attributes [C23.550.291]
16. Dysbiosis [C23.550.308]
17. Emphysema [C23.550.325]
18. Extravasation of Diagnostic and Therapeutic Materials [C23.550.340]
19. Femoracetabular Impingement [C23.550.347]
20. Fibrosis [C23.550.355]
21. Frailty [C23.550.359]
22. Genomic Instability [C23.550.362]
23. Gliosis [C23.550.369]
24. Granuloma [C23.550.382]
25. Granulomatosis, Orofacial [C23.550.384]
26. Growth Disorders [C23.550.393]
27. Hemolysis [C23.550.403]
28. Hemorrhage [C23.550.414]
29. Hyperammonemia [C23.550.421]
30. Hyperamylasemia [C23.550.425]
31. Hyperbilirubinemia [C23.550.429]
32. Hyperplasia [C23.550.444]
33. Hyperuricemia [C23.550.449]
34. Hypovolemia [C23.550.455]
35. Inflammation [C23.550.470]
36. Intraoperative Complications [C23.550.505]
37. Ischemia [C23.550.513]
38. Leukoaraiosis [C23.550.522]
39. Leukocytosis [C23.550.526]
40. Lithiasis [C23.550.537]
41. Long Term Adverse Effects [C23.550.543]
42. Malacoplakia [C23.550.548]
43. Menstruation Disturbances [C23.550.568]
44. Metaplasia [C23.550.589]
45. Muscle Weakness [C23.550.695]
46. Myotoxicity [C23.550.706]
47. Necrosis [C23.550.717]
48. Neointima [C23.550.722]
49. Neoplastic Processes [C23.550.727]
50. Nerve Degeneration [C23.550.737]
51. Ochronosis [C23.550.744]
52. Ossification, Heterotopic [C23.550.751]
53. Ototoxicity [C23.550.753]
54. Pigmentation Disorders [C23.550.755]
55. Polydipsia [C23.550.759]
56. Postoperative Complications [C23.550.767]
57. Afferent Loop Syndrome [C23.550.767.050]
58. Anastomotic Leak [C23.550.767.071]
59. Breast Cancer Lymphedema [C23.550.767.082]
60. Corneal Endothelial Cell Loss [C23.550.767.093]
61. Coronary-Subclavian Steal Syndrome [C23.550.767.115]
62. Delayed Emergence from Anesthesia [C23.550.767.137]
63. Emergence Delirium [C23.550.767.181]
64. Failed Back Surgery Syndrome [C23.550.767.225]
65. Graft Occlusion, Vascular [C23.550.767.400]
66. Incisional Hernia [C23.550.767.500]
67. Low Anterior Resection Syndrome [C23.550.767.550]
68. Malignant Hyperthermia [C23.550.767.600]
69. Pain, Postoperative [C23.550.767.700]
70. Phantom Limb [C23.550.767.700.500]
71. Postcholecystectomy Syndrome [C23.550.767.775]
72. Postgastrectomy Syndromes [C23.550.767.812]
73. Dumping Syndrome [C23.550.767.812.500]
74. Postoperative Cognitive Complications [C23.550.767.831]
75. Postoperative Hemorrhage [C23.550.767.850]
76. Endoleak [C23.550.767.850.500]
77. Postoperative Nausea and Vomiting [C23.550.767.859]
78. Postpericardiotomy Syndrome [C23.550.767.863]
79. Prosthesis Failure [C23.550.767.865]
80. Implant Capsular Contracture [C23.550.767.865.500]
81. Prosthesis-Related Infections [C23.550.767.868]
82. Reperfusion Injury [C23.550.767.877]
83. Myocardial Reperfusion Injury [C23.550.767.877.500]
84. Post-Cardiac Arrest Syndrome [C23.550.767.877.625]
85. Primary Graft Dysfunction [C23.550.767.877.750]
86. Shock, Surgical [C23.550.767.879]
87. Short Bowel Syndrome [C23.550.767.882]
88. Slit Ventricle Syndrome [C23.550.767.884]
89. Surgical Wound Dehiscence [C23.550.767.887]
90. Surgical Wound Infection [C23.550.767.925]
91. Vasoplegia [C23.550.767.962]

**Intraoperative Complications [C23.550.505]**

1. Blood Loss, Surgical [C23.550.505.300]
2. Intraoperative Awareness [C23.550.505.400]
3. Malignant Hyperthermia [C23.550.505.700]
4. Ischemia [C23.550.513]
5. Chronic Limb-Threatening Ischemia [C23.550.513.178]
6. Infarction [C23.550.513.355]
7. Brain Infarction [C23.550.513.355.250]
8. Hepatic Infarction [C23.550.513.355.500]
9. Myocardial Infarction [C23.550.513.355.750]
10. Pulmonary Infarction [C23.550.513.355.875]
11. Splenic Infarction [C23.550.513.355.937]
12. No-Reflow Phenomenon [C23.550.513.677]
13. Leukoaraiosis [C23.550.522]
14. Leukocytosis [C23.550.526]
15. Lithiasis [C23.550.537]
16. Long-Term Adverse Effects [C23.550.543]
17. Malacoplakia [C23.550.548]
18. Menstruation Disturbances [C23.550.568]
19. Amenorrhea [C23.550.568.500]
20. Dysmenorrhea [C23.550.568.750]
21. Menorrhagia [C23.550.568.875]
22. Oligomenorrhea [C23.550.568.937]
23. Premenstrual Syndrome [C23.550.568.968]
24. Premenstrual Dysphoric Disorder [C23.550.568.968.500]
25. Metaplasia [C23.550.589]
26. Neovascularization, Pathologic [C23.550.589.500]
27. Muscle Weakness [C23.550.695]
28. Dropped Head Syndrome [C23.550.695.500]
29. Myotoxicity [C23.550.706]
30. Necrosis [C23.550.717]
31. Dental Pulp Necrosis [C23.550.717.182]
32. DNA Degradation, Necrotic [C23.550.717.273]
33. Fat Necrosis [C23.550.717.365]
34. Gangrene [C23.550.717.427]
35. Infarction [C23.550.717.489]
36. Brain Infarction [C23.550.717.489.250]
37. Hepatic Infarction [C23.550.717.489.500]
38. Myocardial Infarction [C23.550.717.489.750]
39. Pulmonary Infarction [C23.550.717.489.875]
40. Splenic Infarction [C23.550.717.489.937]
41. Osteonecrosis [C23.550.717.732]
42. Bisphosphonate-Associated Osteonecrosis of the Jaw [C23.550.717.732.183]
43. Femur Head Necrosis [C23.550.717.732.368]
44. Neointima [C23.550.722]
45. Neoplastic Processes [C23.550.727]
46. Anaplasia [C23.550.727.045]
47. Carcinogenesis [C23.550.727.098]
48. Neoplasm Invasiveness [C23.550.727.645]
49. Neoplasm Metastasis [C23.550.727.650]
50. Neoplasm Recurrence, Local [C23.550.727.655]
51. Neoplasm Regression, Spontaneous [C23.550.727.670]
52. Neoplasm, Residual [C23.550.727.700]
53. Oncogene Addiction [C23.550.727.850]
54. Nerve Degeneration [C23.550.737]
55. Retrograde Degeneration [C23.550.737.500]
56. Subacute Combined Degeneration [C23.550.737.625]
57. Wallerian Degeneration [C23.550.737.750]
58. Ochronosis [C23.550.744]
59. Ossification, Heterotopic [C23.550.751]
60. Ossification of Posterior Longitudinal Ligament [C23.550.751.500]
61. Ototoxicity [C23.550.753]
62. Pigmentation Disorders [C23.550.755]
63. Cronkhite-Canada Syndrome [C23.550.755.500]
64. Polydipsia [C23.550.759]
65. Polydipsia, Psychogenic [C23.550.759.500]

**C category Infections** [C01]:

1. Aneurysm, Infected [C01.069]
2. Arthritis, Infectious [C01.100]
3. Asymptomatic Infections [C01.125]
4. Bacterial Infections and Mycoses [C01.150]
5. Bone Diseases, Infectious [C01.160]
6. Breakthrough Infections [C01.175]
7. Cardiovascular Infections [C01.190]
8. Catheter-Related Infections [C01.195]
9. Central Nervous System Infections [C01.207]
10. Coinfection [C01.218]
11. Communicable Diseases [C01.221]
12. Community-Acquired Infections [C01.234]
13. Cross Infection [C01.248]
14. Eye Infections [C01.375]
15. Focal Infection [C01.392]
16. Gingivitis [C01.408]
17. Hepatitis, Animal [C01.436]
18. Intraabdominal Infections [C01.463]
19. Laboratory Infection [C01.503]
20. Latent Infection [C01.550]
21. Opportunistic Infections [C01.597]
22. Parasitic Diseases [C01.610]
23. Pelvic Infection [C01.635]
24. Persistent Infection [C01.645]
25. Poult Enteritis Mortality Syndrome [C01.655]
26. Pregnancy Complications, Infectious [C01.674]
27. Prosthesis-Related Infections [C01.685]
28. Reproductive Tract Infections [C01.730]
29. Respiratory Tract Infections [C01.748]
30. Sepsis [C01.757]
31. Sexually Transmitted Diseases [C01.778]
32. Skin Diseases, Infectious [C01.800]
33. Soft Tissue Infections [C01.820]
34. Suppuration [C01.830]
35. Toxemia [C01.861]
36. Urinary Tract Infections [C01.915]
37. Vaccine-Preventable Diseases [C01.918]
38. Vector Borne Diseases [C01.920]
39. Virus Diseases [C01.925]
40. Waterborne Diseases [C01.936]
41. Wound Infection [C01.947]
42. Zoonoses [C01.973]
43. Neoplasms [C04]
44. Cysts [C04.182]
45. Hamartoma [C04.445]
46. Neoplasms by Histologic Type [C04.557]
47. Neoplasms by Site [C04.588]
48. Neoplasms, Experimental [C04.619]
49. Neoplasms, Hormone-Dependent [C04.626]
50. Neoplasms, Multiple Primary [C04.651]
51. Neoplasms, Post-Traumatic [C04.666]
52. Neoplasms, Radiation-Induced [C04.682]
53. Neoplasms, Second Primary [C04.692]
54. Neoplastic Processes [C04.697]
55. Neoplastic Syndromes, Hereditary [C04.700]
56. Paraneoplastic Syndromes [C04.730]
57. Precancerous Conditions [C04.834]
58. Pregnancy Complications, Neoplastic [C04.850]
59. Musculoskeletal Diseases [C05]
60. Bone Diseases [C05.116]
61. Cartilage Diseases [C05.182]
62. Fasciitis [C05.321]
63. Foot Deformities [C05.330]
64. Foot Diseases [C05.360]
65. Hand Deformities [C05.390]
66. Jaw Diseases [C05.500]
67. Joint Diseases [C05.550]
68. Muscular Diseases [C05.651]
69. Musculoskeletal Abnormalities [C05.660]
70. Rheumatic Diseases [C05.799]
71. Digestive System Diseases [C06]
72. Biliary Tract Diseases [C06.130]
73. Digestive System Abnormalities [C06.198]
74. Digestive System Fistula [C06.267]
75. Digestive System Neoplasms [C06.301]
76. Gastrointestinal Diseases [C06.405]
77. Liver Diseases [C06.552]
78. Pancreatic Diseases [C06.689]
79. Peritoneal Diseases [C06.844]
80. Stomatognathic Diseases [C07]
81. Ankyloglossia [C07.160]
82. Jaw Diseases [C07.320]
83. Mouth Diseases [C07.465]
84. Pharyngeal Diseases [C07.550]
85. Stomatognathic System Abnormalities [C07.650]
86. Temporomandibular Joint Disorders [C07.678]
87. Tooth Diseases [C07.793]
88. Respiratory Tract Diseases [C08]
89. Bronchial Diseases [C08.127]
90. Ciliary Motility Disorders [C08.200]
91. Granuloma, Respiratory Tract [C08.280]
92. Laryngeal Diseases [C08.360]
93. Lung Diseases [C08.381]
94. Nose Diseases [C08.460]
95. Pleural Diseases [C08.528]
96. Respiration Disorders [C08.618]
97. Respiratory Hypersensitivity [C08.674]
98. Respiratory System Abnormalities [C08.695]
99. Respiratory Tract Fistula [C08.702]
100. Respiratory Tract Infections [C08.730]
101. Respiratory Tract Neoplasms [C08.785]
102. Thoracic Diseases [C08.846]
103. Tracheal Diseases [C08.907]
104. Otorhinolaryngologic Diseases [C09]
105. Ciliary Motility Disorders [C09.150]
106. Ear Diseases [C09.218]
107. Laryngeal Diseases [C09.400]
108. Nose Diseases [C09.603]
109. Otorhinolaryngologic Neoplasms [C09.647]
110. Pharyngeal Diseases [C09.775]
111. Nervous System Diseases [C10]
112. Autoimmune Diseases of the Nervous System [C10.114]
113. Autonomic Nervous System Diseases [C10.177]
114. Central Nervous System Diseases [C10.228]
115. Chronobiology Disorders [C10.281]
116. Cranial Nerve Diseases [C10.292]
117. Demyelinating Diseases [C10.314]
118. Nervous System Malformations [C10.500]
119. Nervous System Neoplasms [C10.551]
120. Neurocutaneous Syndromes [C10.562]
121. Neurodegenerative Diseases [C10.574]
122. Neuroinflammatory Diseases [C10.586]
123. Neurologic Manifestations [C10.597]
124. Neuromuscular Diseases [C10.668]
125. Neurotoxicity Syndromes [C10.720]
126. Restless Legs Syndrome [C10.803]
127. Sleep Wake Disorders [C10.886]
128. Trauma, Nervous System [C10.900]
129. Eye Diseases [C11]
130. Asthenopia [C11.093]
131. Cogan Syndrome [C11.180]
132. Conjunctival Diseases [C11.187]
133. Corneal Diseases [C11.204]
134. Eye Abnormalities [C11.250]
135. Eye Diseases, Hereditary [C11.270]
136. Eye Hemorrhage [C11.290]
137. Eye Infections [C11.294]
138. Eye Injuries [C11.297]
139. Eye Manifestations [C11.300]
140. Eye Neoplasms [C11.319]
141. Eyelid Diseases [C11.338]
142. Lacrimal Apparatus Diseases [C11.496]
143. Lens Diseases [C11.510]
144. Ocular Hypertension [C11.525]
145. Ocular Hypotension [C11.540]
146. Ocular Motility Disorders [C11.590]
147. Optic Nerve Diseases [C11.640]
148. Orbital Diseases [C11.675]
149. Pupil Disorders [C11.710]
150. Refractive Errors [C11.744]
151. Retinal Diseases [C11.768]
152. Scleral Diseases [C11.790]
153. Uveal Diseases [C11.941]
154. Vision Disorders [C11.966]
155. Vitreous Detachment [C11.980]
156. Urogenital Diseases [C12]
157. Female Urogenital Diseases and Pregnancy Complications [C12.050]
158. Genital Diseases [C12.100]
159. Male Urogenital Diseases [C12.200]
160. Pelvic Floor Disorders [C12.400]
161. Tuberculosis, Urogenital [C12.600]
162. Urogenital Abnormalities [C12.800]
163. Urogenital Neoplasms [C12.900]
164. Urologic Diseases [C12.950]
165. Cardiovascular Diseases [C14]
166. Cardiovascular Abnormalities [C14.240]
167. Cardiovascular Infections [C14.260]
168. Heart Diseases [C14.280]
169. Pregnancy Complications, Cardiovascular [C14.583]
170. Vascular Diseases [C14.907]
171. Hemic and Lymphatic Diseases [C15]
172. Hematologic Diseases [C15.378]
173. Lymphatic Diseases [C15.604]
174. Congenital, Hereditary, and Neonatal Diseases and Abnormalities [C16]
175. Congenital Abnormalities [C16.131]
176. Fetal Diseases [C16.300]
177. Genetic Diseases, Inborn [C16.320]
178. Infant, Newborn, Diseases [C16.614]
179. Skin and Connective Tissue Diseases [C17]
180. Connective Tissue Diseases [C17.300]
181. Skin Diseases [C17.800]
182. Nutritional and Metabolic Diseases [C18]
183. Metabolic Diseases [C18.452]
184. Nutrition Disorders [C18.654]
185. Endocrine System Diseases [C19]
186. Adrenal Gland Diseases [C19.053]
187. Bone Diseases, Endocrine [C19.149]
188. Diabetes Mellitus [C19.246]
189. Dwarfism [C19.297]
190. Endocrine Gland Neoplasms [C19.344]
191. Gonadal Disorders [C19.391]
192. Parathyroid Diseases [C19.642]
193. Pituitary Diseases [C19.700]
194. Polyendocrinopathies, Autoimmune [C19.787]
195. Thyroid Diseases [C19.874]
196. Tuberculosis, Endocrine [C19.927]
197. Immune System Diseases [C20]
198. Autoimmune Diseases [C20.111]
199. Erythroblastosis, Fetal [C20.306]
200. Glomerulonephritis, Membranoproliferative [C20.425]
201. Graft vs Host Disease [C20.452]
202. Hypersensitivity [C20.543]
203. Immune Reconstitution Inflammatory Syndrome [C20.608]
204. Immunologic Deficiency Syndromes [C20.673]
205. Immunoproliferative Disorders [C20.683]
206. Mast Cell Activation Disorders [C20.762]
207. Purpura, Thrombocytopenic [C20.841]
208. Transfusion Reaction [C20.920]
209. Disorders of Environmental Origin [C21]
210. Environmental Illness [C21.223]
211. Preconception Injuries [C21.676]
212. Animal Diseases [C22]
213. Pathological Conditions, Signs and Symptoms [C23]
214. Morphological and Microscopic Findings [C23.149]
215. Pathological Conditions, Anatomical [C23.300]
216. Pathologic Processes [C23.550]
217. Signs and Symptoms [C23.888]
218. Occupational Diseases [C24]
219. Chemically-Induced Disorders [C25]
220. Wounds and Injuries [C26]
221. Abdominal Injuries [C26.017]
222. Accidental Injuries [C26.040]
223. Amputation, Traumatic [C26.062]
224. Arm Injuries [C26.088]
225. Asphyxia [C26.103]
226. Athletic Injuries [C26.115]
227. Back Injuries [C26.117]
228. Barotrauma [C26.120]
229. Birth Injuries [C26.141]
230. Bites and Stings [C26.176]
231. Burns [C26.200]
232. Cold Injury [C26.212]
233. Contrecoup Injury [C26.224]
234. Crush Injuries [C26.257]
235. Joint Dislocations [C26.289]
236. Drowning [C26.304]
237. Electric Injuries [C26.324]
238. Esophageal Perforation [C26.348]
239. Extravasation of Diagnostic and Therapeutic Materials [C26.371]
240. Foreign Bodies [C26.392]
241. Fractures, Bone [C26.404]
242. Fractures, Cartilage [C26.411]
243. Frostbite [C26.417]
244. Hand Injuries [C26.448]
245. Heat Stress Disorders [C26.522]
246. Hip Injuries [C26.531]
247. Lacerations [C26.540]
248. Leg Injuries [C26.558]
249. Microtrauma, Physical [C26.599]
250. Multiple Trauma [C26.640]
251. Nasal Septal Perforation [C26.670]
252. Neck Injuries [C26.700]
253. Occupational Injuries [C26.716]
254. Radiation Injuries [C26.733]
255. Reinjuries [C26.741]
256. Retropneumoperitoneum [C26.748]
257. Rupture [C26.761]
258. Self Mutilation [C26.780]
259. Shock, Traumatic [C26.797]
260. Shoulder Injuries [C26.803]
261. Soft Tissue Injuries [C26.808]
262. Spinal Cord Injuries [C26.819]
263. Sprains and Strains [C26.844]
264. Surgical Wound [C26.859]
265. Tendon Injuries [C26.874]
266. Thoracic Injuries [C26.891]
267. Tooth Injuries [C26.900]
268. Trauma, Nervous System [C26.915]
269. Tympanic Membrane Perforation [C26.930]
270. Vascular System Injuries [C26.940]
271. War-Related Injuries [C26.946]
272. Wounds, Nonpenetrating [C26.974]
273. Wounds, Penetrating [C26.986]

**Prompt 2.**

**Identifying the Text Preceding a Surgical Intervention**

You are given a patient’s description and a surgical intervention performed on the patient.

The task is to extract the chronological narrative of events that occurred **up to but not including** the surgical intervention.

- Do not include the description of the surgical procedure itself.
- End the extracted text immediately **before the first mention of the surgical intervention**.
- Ensure the output contains only events that preceded the surgery.

**Output:** The text preceding the surgical intervention (ending exactly before the surgery is mentioned).

**Input:**
Patient’s description: [Insert full case text]
Surgical Intervention: [Insert surgical procedure]

\###

Example:

**Input:**

**The Patient’s description:**

Maria G., a 74-year-old female, presented to the emergency department with a 2-day history of severe epigastric pain radiating to the back, associated with nausea and multiple episodes of non-bloody vomiting. She reported decreased appetite and subjective fever. Her past medical history included gallstones, hyperlipidemia, and coronary artery disease. On physical examination, the patient was febrile (38.7°C), with tenderness in the right upper quadrant and epigastrium. Laboratory findings revealed elevated white blood cell count, increased serum lipase (1120 U/L), and mildly abnormal liver function tests. Abdominal ultrasound showed gallstones without ductal dilatation, while CT imaging confirmed acute gallstone pancreatitis, with signs of peripancreatic fat stranding and inflammation. The patient was admitted and initially managed with intravenous fluids, analgesia, and bowel rest. On hospital day 3, due to persistent symptoms and recurrent pain, the surgical team proceeded with a laparoscopic cholecystectomy. Intraoperative findings included an inflamed gallbladder with multiple small stones. The surgery was uneventful. However, on postoperative day 2, the patient developed fever (39.1°C), tachycardia, and mild hypotension. Repeat labs showed worsening inflammatory markers. A CT scan of the abdomen revealed a small fluid collection in the gallbladder fossa, consistent with a postoperative localized intra-abdominal infection. The patient was managed conservatively with intravenous antibiotics (piperacillin-tazobactam), and no drainage procedure was required. She gradually improved and was discharged on postoperative day 7.

The surgical intervention: **laparoscopic cholecystectomy**

**###**

**Output:**

Maria G., a 74-year-old female, presented to the emergency department with a 2-day history of severe epigastric pain radiating to the back, associated with nausea and multiple episodes of non-bloody vomiting. She reported decreased appetite and subjective fever. Her past medical history included gallstones, hyperlipidemia, and coronary artery disease. On physical examination, the patient was febrile (38.7°C), with tenderness in the right upper quadrant and epigastrium. Laboratory findings revealed elevated white blood cell count, increased serum lipase (1120 U/L), and mildly abnormal liver function tests. Abdominal ultrasound showed gallstones without ductal dilatation, while CT imaging confirmed acute gallstone pancreatitis, with signs of peripancreatic fat stranding and inflammation. The patient was admitted and initially managed with intravenous fluids, analgesia, and bowel rest. On hospital day 3, due to persistent symptoms and recurrent pain, the surgical team proceeded with—

**Prompt 3**

**Identifying Risk Factors that might Lead to Surgical Complications**

**Definition:**
A *risk factor* is any patient-related, disease-related, anatomical, physiological, or procedural characteristic present *before* the surgical intervention that increases the likelihood of **intraoperative or postoperative surgical complications** (NOT general adverse outcomes).

**Task:** You are an expert surgeon and clinical risk analysis. Given a medical case text, the patient’s DIAGNOSIS and a specified SURGICAL INTERVENTION, your task is to identify and extract all **clinically relevant preoperative risk factors** that have **established, strong correlations** with surgical complications from that intervention.

**Input:**

- Medical Case Text: {Insert case text here}
- DIAGNOSIS: {DIAGNOSIS HERE}
- SURGICAL INTERVENTION: {SURGICAL INTERVENTION HERE}

**Requirements:**

1. Focus specifically on risk factors for **surgical complications,** including:
   - Intraoperative complications (bleeding, organ injury, conversion to open surgery, anesthetic complications)
   - Postoperative complications (wound infection, bile leaks, bleeding, pneumonia, thromboembolism, delayed recovery)
   - NOT general medical outcomes or mortality risk
2. Only include risk factors with **established, strong clinical evidence** for increasing surgical complication risk:
   - Must have documented correlation in surgical literature
   - Exclude weak, questionable, or indirect associations
   - Exclude factors that primarily affect general medical outcomes rather than surgical complications
3. Extract relevant information, including:
   - Patient demographics (age, BMI if available) - only if strongly correlated
   - Comorbidities with established surgical risk (e.g., active infection, severe cardiac disease, coagulopathy)
   - Anatomical and pathological conditions affecting surgical difficulty (e.g., active inflammation, anatomical distortion)
   - Laboratory abnormalities directly affecting surgery (e.g., coagulation defects, severe organ dysfunction)
   - Imaging findings indicating surgical complexity
   - Clinical status indicators affecting surgical risk (e.g., hemodynamic instability, active infection)
4. Classify each risk factor by type:
   - Demographic, Comorbidity, Anatomical/Pathological, Laboratory/Physiological, Procedural history, Clinical status
5. Perform internal quality control to ensure extracted risk factors:
   - Have a **strong, established** correlation with surgical complications
   - Are specific to the surgical intervention being performed
   - Directly impact intraoperative or postoperative surgical outcomes

**Output Format:** Provide only the final list of extracted risk factors in this exact format: [Risk Factor ICD-10-CM Name, Risk Factor Type]

**Notes:**

- Include only risk factors with robust clinical evidence for surgical complication correlation
- Exclude factors with weak, questionable, or primarily non-surgical outcome associations
- Provide no additional commentary or explanation beyond the required output format

**Example:**

**Input:**

Medical Case Text: {Maria G., a 74-year-old female, presented to the emergency department with a 2-day history of severe epigastric pain radiating to the back, associated with nausea and multiple episodes of non-bloody vomiting. She reported decreased appetite and subjective fever. Her past medical history included gallstones, hyperlipidemia, and coronary artery disease. On physical examination, the patient was febrile (38.7°C), with tenderness in the right upper quadrant and epigastrium. Laboratory findings revealed elevated white blood cell count, increased serum lipase (1120 U/L), and mildly abnormal liver function tests. Abdominal ultrasound showed gallstones without ductal dilatation, while CT imaging confirmed acute gallstone pancreatitis, with signs of peripancreatic fat stranding and inflammation. The patient was admitted and initially managed with intravenous fluids, analgesia, and bowel rest. On hospital day 3, due to persistent symptoms and recurrent pain, the surgical team proceeded with—}

DIAGNOSIS: {**Gallstone Pancreatitis**}

SURGICAL INTERVENTION: {**laparoscopic cholecystectomy**}

**Output:**

**RISK FACTORS:**

[Age 74 years, Demographic]

[Acute pancreatitis, Anatomical/Pathological]

[Elevated white blood cell count, Laboratory/Physiological]

[Peripancreatic inflammation, Anatomical/Pathological]

Prompt 4A

________________________________________

Virtual Patients Simulation

________________________________________

**Introduction:**

- A virtual patient is a representation of a patient’s clinical profile

with the purpose of simulating and predicting health outcomes.

- A **patient’s profile** includes his/her (1) DIAGNOSIS, (2) planned SURGICAL INTERVENTION, and (3) RISK FACTORS.
- Your task is to generate a Virtual Patients Cohort from a patient’s profile.

**The structure of the prompt:**

- **Module 1** determines the data type of each RISK FACTOR.
- **Module 2** identifies the distribution of each RISK FACTOR.
- **Module 3** generates up to 50 unique virtual patients from the patient's profile.

**Your input is:**

(1) The patient's RISK FACTORS: [PATIENT’S risk factors HERE]
(2) The patient’s DIAGNOSIS: [DIAGNOSIS HERE]
(3) The planned SURGICAL INTERVENTION: [SURGICAL INTERVENTION HERE]
(4) TIMING of the complication: [TIMING HERE]

**Note**: The TIMING of a complication corresponds with the following typology:

**1. Immediate Complications**

- **Timing:** During surgery or within **24 hours**
- **Examples:**
  - Hemorrhage (intraoperative or postoperative bleeding)
  - Anesthesia-related issues (e.g., aspiration, cardiac events)
  - Injury to adjacent structures
  - Acute allergic reactions

**2. Early Complications**

- **Timing:** Within **24 hours to 30 days** after surgery
- **Examples:**
  - Surgical site infection (SSI)
  - Pneumonia
  - Deep vein thrombosis (DVT) or pulmonary embolism (PE)
  - Paralytic ileus or bowel obstruction
  - Urinary leakage or fistula
  - Acute kidney injury
  - Wound dehiscence

**3. Late Complications**

- **Timing:** After **30 days**, often up to **months post-op**
- **Examples:**
  - Anastomotic strictures
  - Hernia at the surgical site
  - Ureteral obstruction or stricture
  - Chronic pain
  - Delayed infections (e.g., deep abscess)

**4. Long-term (or Chronic) Complications**

- **Timing:** **Months to years** after surgery
- **Examples:**
  - **Neoplasm recurrence**
  - Chronic kidney disease (from urinary diversion)
  - Metabolic disturbances (e.g., acidosis with ileal conduit)
  - Sexual dysfunction
  - Long-term urinary incontinence or retention

**Final output:**

A list of the virtual patients, each with his/her unique parameters of the risk factors.

Present the output in JSON structure.

___________________________________________________________________

**Internal quality check:**

- Perform an internal quality check
- Ensure you generated DIFFERENT and unique virtual patients from each **patient’s profile**.
- Ensure that each module has been appropriately applied

___________________________________________________________________

**THE MODULES**

**Module 1. Identify the Data Type of Each RISK FACTOR**

**Task.**

Determine the data type for each RISK FACTOR.

The data type can be continuous, binary, or categorical.

o For binary RISK FACTORS (e.g., diabetes mellitus, hypertension), specify the two possible values the RISK FACTOR can take, typically 0 (absence of the RISK FACTOR) and 1 (presence of the RISK FACTOR).

o For categorical RISK FACTORS (e.g., smoking status), specify the possible categories and their corresponding values (e.g., Non-smoker = 0, Light smoker = 1, Heavy smoker = 2).

Examples:

1. Advanced Age

o Data Type: Continuous

o Description: Age in years

2. Gender

o Data Type: Binary

o Values: 0 = Female, 1 = Male

3. Diabetes Mellitus

o Data Type: Binary

o Values: 0 = Absence of diabetes, 1 = Presence of diabetes

________________________________________

**Module 2.** Determine the Appropriate Statistical Distribution for Each RISK FACTOR

**Input:** Use the list of RISK FACTORS and their data types produced by module 1.

**Task:** Determine the Distribution Type for Each RISK FACTOR.

**Subtasks:**

**Subtask 2.1:**

o Identify the most appropriate distribution type for each RISK FACTOR, incorporating general medical knowledge specific to patients diagnosed with [DIAGNOSIS] and planned to have the following surgical intervention: [SURGICAL INTERVENTION].

Use the following guidelines for Choosing Distributions by Data Type:

• Continuous RISK FACTORS: Advise the selection of a distribution based on the data’s natural limits and spread within the target population:

o Normal: When values are symmetrically distributed around a mean.

o Truncated Normal: When values are concentrated near an average but are limited by realistic biological constraints (e.g., age).

o Log-normal: When values are right-skewed, common in medical data for age or time-to-event RISK FACTORS.

• Binary RISK FACTORS: Suggest using Bernoulli distributions, specifying that probability estimates should reflect the prevalence within the patient population.

• Categorical RISK FACTORS: Recommend using a multinomial distribution with proportions based on medical literature or population data if available. For example, “For smoking status, use values based on prevalence among similar patients.”

o Ensure the distribution describes the Probability Density Function (PDF) of the RISK FACTOR.

o If you do not have medical knowledge about the distribution, guess the most likely distribution.

**Subtask 2.2:**

o Select the most appropriate parameters for each distribution you identified in subtask 2.1.

o The parameters must suit the medical diagnosis of [DIAGNOSIS].

o If you have no medical knowledge to support your decision, use parameters that maximize the entropy of the selected distribution.

Output format:

For each RISK FACTOR:

o RISK FACTOR NAME

o DATA TYPE

o DISTRIBUTION

o RANGE OF VALUES

o DISTRIBUTION PARAMETERS

Examples:

1. Advanced Age

o RISK FACTOR in MeSH terms: [Aged, 80 and over]

o Data Type: Continuous

o Distribution: Truncated Normal

o Range of Values: 60 to 100 years (targeting an elderly population)

o Distribution Parameters:

♣ Mean (μ) ≈ 85

♣ Standard deviation (σ) ≈ 5

♣ Lower bound = 60, Upper bound = 100

2. Gender

o RISK FACTOR in MeSH terms: [Male]

o Data Type: Binary

o Distribution: Bernoulli

o Range of Values: 0 (Female), 1 (Male)

o Distribution Parameters:

♣ Probability of Male (p) ≈ 0.6 (assuming slight male predominance in cholangitis cases)

3. Diabetes Mellitus

o RISK FACTOR in MeSH terms: [Diabetes Mellitus]

o Data Type: Binary

o Distribution: Bernoulli

o Range of Values: 0 (No Diabetes), 1 (Presence of Diabetes)

o Distribution Parameters:

♣ Probability of Presence of Diabetes (p) ≈ 0.4 (increased prevalence in elderly populations)

________________________________________

**Module 3. Generating a Cohort of Virtual Patients**

**Objective:**

- Generate a cohort of up to 50 DIFFERENT virtual patients diagnosed with [DIAGNOSIS HERE], and planned to have the surgical intervention of [SURGICAL INTERVENTION HERE].
- Each virtual patient should include values for all specified RISK FACTORS.
- **VERY IMPORTANT:** The DIAGNOSIS and SURGICAL INTERVENTION are constants for each virtual patient. The difference between the virtual patients is in the parameters’ values of each RISK FACTOR.

**INSTRUCTIONS:**

1. Sampling

• Random Sampling:

o For each RISK FACTOR, sample a value according to its specified distribution type (e.g., Normal, Bernoulli), range, and parameters.

o Ensure sampling reflects realistic distributions coherent with the described [DIAGNOSIS], incorporating variations around the baseline clinical case.

• Clinical Plausibility:

o Check each generated virtual patient’s sampled values for clinical plausibility. For instance, ensure advanced age aligns with comorbidities like hypertension or ischemic heart disease.

o Resample any implausible combinations to maintain coherence with known profiles of patients suffering from [DIAGNOSIS].

• Parameter Correlations:

o Model known correlations between RISK FACTORS (e.g., advanced age with hypertension, obesity with diabetes), especially when these correlations contribute to a higher likelihood of surgical complications.

________________________________________

2. Unique Combination Requirement

- Each virtual patient must express a **unique combination** of RISK FACTOR values within plausible ranges.
- The cohort can include **fewer than 50 patients** if fewer than 50 clinically plausible unique combinations exist.
- Ensure no duplicates across all RISK FACTOR values in the cohort.

________________________________________

**Example**

**Input:**

(1) The patient's RISK FACTORS:

[Age 74 years, Demographic]

[Acute pancreatitis, Anatomical/Pathological]

[Elevated white blood cell count, Laboratory/Physiological]

[Peripancreatic inflammation, Anatomical/Pathological]

(2) The patient’s DIAGNOSIS: **Gallstone Pancreatitis**
(3) The planned SURGICAL INTERVENTION: **laparoscopic cholecystectomy**
(4) TIMING: IMMEDIATE OR EARLY.

**Output example of first ten virtual patients:**

[

{

"VirtualPatientID": "VP1",

"DIAGNOSIS": "Gallstone Pancreatitis",

"SURGICAL_INTERVENTION": "Laparoscopic Cholecystectomy",

"RISK_FACTORS": {

"Age": 72,

"Acute pancreatitis": 1,

"Elevated white blood cell count": 15,

"Peripancreatic inflammation": 1

}

},

{

"VirtualPatientID": "VP2",

"DIAGNOSIS": "Gallstone Pancreatitis",

"SURGICAL_INTERVENTION": "Laparoscopic Cholecystectomy",

"RISK_FACTORS": {

"Age": 77,

"Acute pancreatitis": 1,

"Elevated white blood cell count": 12,

"Peripancreatic inflammation": 1

}

},

{

"VirtualPatientID": "VP3",

"DIAGNOSIS": "Gallstone Pancreatitis",

"SURGICAL_INTERVENTION": "Laparoscopic Cholecystectomy",

"RISK_FACTORS": {

"Age": 69,

"Acute pancreatitis": 1,

"Elevated white blood cell count": 18,

"Peripancreatic inflammation": 1

}

},

{

"VirtualPatientID": "VP4",

"DIAGNOSIS": "Gallstone Pancreatitis",

"SURGICAL_INTERVENTION": "Laparoscopic Cholecystectomy",

"RISK_FACTORS": {

"Age": 75,

"Acute pancreatitis": 1,

"Elevated white blood cell count": 16,

"Peripancreatic inflammation": 1

}

},

{

"VirtualPatientID": "VP5",

"DIAGNOSIS": "Gallstone Pancreatitis",

"SURGICAL_INTERVENTION": "Laparoscopic Cholecystectomy",

"RISK_FACTORS": {

"Age": 70,

"Acute pancreatitis": 1,

"Elevated white blood cell count": 13,

"Peripancreatic inflammation": 1

}

},

{

"VirtualPatientID": "VP6",

"DIAGNOSIS": "Gallstone Pancreatitis",

"SURGICAL_INTERVENTION": "Laparoscopic Cholecystectomy",

"RISK_FACTORS": {

"Age": 74,

"Acute pancreatitis": 1,

"Elevated white blood cell count": 14,

"Peripancreatic inflammation": 1

}

},

{

"VirtualPatientID": "VP7",

"DIAGNOSIS": "Gallstone Pancreatitis",

"SURGICAL_INTERVENTION": "Laparoscopic Cholecystectomy",

"RISK_FACTORS": {

"Age": 71,

"Acute pancreatitis": 1,

"Elevated white blood cell count": 17,

"Peripancreatic inflammation": 1

}

},

{

"VirtualPatientID": "VP8",

"DIAGNOSIS": "Gallstone Pancreatitis",

"SURGICAL_INTERVENTION": "Laparoscopic Cholecystectomy",

"RISK_FACTORS": {

"Age": 73,

"Acute pancreatitis": 1,

"Elevated white blood cell count": 11,

"Peripancreatic inflammation": 1

}

},

{

"VirtualPatientID": "VP9",

"DIAGNOSIS": "Gallstone Pancreatitis",

"SURGICAL_INTERVENTION": "Laparoscopic Cholecystectomy",

"RISK_FACTORS": {

"Age": 76,

"Acute pancreatitis": 1,

"Elevated white blood cell count": 16,

"Peripancreatic inflammation": 1

}

},

{

"VirtualPatientID": "VP10",

"DIAGNOSIS": "Gallstone Pancreatitis",

"SURGICAL_INTERVENTION": "Laparoscopic Cholecystectomy",

"RISK_FACTORS": {

"Age": 68,

"Acute pancreatitis": 1,

"Elevated white blood cell count": 19,

"Peripancreatic inflammation": 1

}

}

]

Prompt 4B

**Prediction of Complications**

**Input:**

The JSON array of virtual patients, where each patient has a unique profile of risk factors (e.g., AcutePancreatitis, WBC, PeripancreaticInflammation).

All the virtual patients have the same:

(1) DIAGNOSIS: [DIAGNOSIS HERE]
(2) PLANNED SURGICAL INTERVENTION: [PLANNED SURGICAL INTERVENTION HERE]
(3) TIMING: [TIMING HERE]

The JSON array is: [JSON ARRAY HERE]

**Task Objective:**
For EACH virtual patient in the JSON array, use

(1) the unique profile of RISK FACTORS and

(2) DIAGNOSIS, PLANNED SURGICAL INTERVENTION, and TIMING to predict the 10 most likely [TIMING HERE] surgical complications.

**Instructions for Processing the JSON Input:**

1. Read each virtual patient’s profile from the JSON input.
2. Consider all available risk factors to identify complications **causally related to the planned surgical intervention**.
3. Use the most updated clinical knowledge for complication prediction.
4. Focus strictly on **surgical complications**—do not include general adverse outcomes or unrelated medical events.
5. For each predicted complication, identify the **most specific MeSH descriptor and MeSH tree number** from the provided <MeSH Tree Data>.
6. Use <Identifying the Specific MeSH Descriptor and Tree Number for a Surgical Complication> as a guide for identifying the MeSH descriptor and tree number.
7. Ensure that the MeSH tree number is **as specific as possible** in the hierarchy. Avoid returning general categories like “postoperative complications” [C23.550.767] or “intraoperative complications” [C23.550.505].
8. Ensure you predict ten complications for each of the given virtual patients.

**Output Format (JSON):**
For each patient, return a JSON object with:

{

"VirtualPatientID": "<VP ID>",

"PredictedComplications": [

{

"ComplicationName": "<MeSH Descriptor>",

"MeSHTreeNumber": "<Tree Number>",

"NormalizedProbability": "<Probability Across the VPs>"

},

...

(10 complications per patient)

]

}

\###

**<Identifying the Specific MeSH Descriptor and Tree Number for a Surgical Complication>**

**Task Objective:**

**For EACH of the 10 predicted complications:**

Identify the most specific **MeSH Descriptor and tree number** that best describes each complication.

**Process:**

8. **Understand the Relationship Between the Diagnosis, the Surgical Intervention, and the Predicted Complication:**
   - Each surgical intervention may have specific complications associated with it.
   - For each complication, a corresponding MeSH tree number describes it.
   - The **MeSH tree number** hierarchy organizes medical terms, so the more specific the complication, the deeper it is in the hierarchy.
9. **Use the <MeSH Tree Data>**
10. **Determine the Most Specific MeSH Tree Number:**
    - Find the complication with the most specific **MeSH tree number**.
    - Return ONLY a single tree number.
    - Use clinical medical knowledge to ensure the MeSH tree number is the most specific and accurate representation of the given surgical complication.
    - Return a dictionary with each **complication descriptor** as the key and its **MeSH tree number** as the value.
11. **IMPORTANT:**

- NEVER return the MeSH tree numbers corresponding to the general **descriptors** "postoperative complications" or "intraoperative complications".
- If the most accurate and specific MeSH tree number does **not** appear in the **<MeSH Tree Data>,** locate it in the **broader MeSH hierarchy** (outside the given subset).
- Focus only on the MeSH tree categories C (i.e., Diseases) and E (i.e., Analytic, Diagnostic, and Therapeutic Techniques)

12. **Error Handling:**
    - If a complication is not found under the specified intervention, return a message saying no matching complications were found.

**Example:**

**Input:**

**Surgical Intervention**: [Laparoscopic Cholecystectomy]

**MeSH Complication Descriptor**: [Intra-abdominal infection (localized fluid collection in gallbladder fossa)]

**Output:**

[Abdominal Abscess, C01.830.025.020]

**Notes:**

- The **MeSH tree number** should be chosen based on clinical knowledge and the hierarchy of terms in MeSH.
- If multiple descriptors describe a complication, use the most specific tree number.

\###

**<MeSH Tree Data>**

Postoperative complications MeSH

92. Pathological Conditions, Signs and Symptoms [C23]
93. Pathologic Processes [C23.550]
94. Acantholysis [C23.550.035]
95. Arrhythmias, Cardiac [C23.550.073]
96. Ascites [C23.550.081]
97. Atrial Remodeling [C23.550.113]
98. Azotemia [C23.550.145]
99. Cardiotoxicity [C23.550.161]
100. Channelopathies [C23.550.177]
101. Chromosome Aberrations [C23.550.210]
102. Death [C23.550.260]
103. Dehydration [C23.550.274]
104. Delayed Graft Function [C23.550.277]
105. Disease [C23.550.288]
106. Disease Attributes [C23.550.291]
107. Dysbiosis [C23.550.308]
108. Emphysema [C23.550.325]
109. Extravasation of Diagnostic and Therapeutic Materials [C23.550.340]
110. Femoracetabular Impingement [C23.550.347]
111. Fibrosis [C23.550.355]
112. Frailty [C23.550.359]
113. Genomic Instability [C23.550.362]
114. Gliosis [C23.550.369]
115. Granuloma [C23.550.382]
116. Granulomatosis, Orofacial [C23.550.384]
117. Growth Disorders [C23.550.393]
118. Hemolysis [C23.550.403]
119. Hemorrhage [C23.550.414]
120. Hyperammonemia [C23.550.421]
121. Hyperamylasemia [C23.550.425]
122. Hyperbilirubinemia [C23.550.429]
123. Hyperplasia [C23.550.444]
124. Hyperuricemia [C23.550.449]
125. Hypovolemia [C23.550.455]
126. Inflammation [C23.550.470]
127. Intraoperative Complications [C23.550.505]
128. Ischemia [C23.550.513]
129. Leukoaraiosis [C23.550.522]
130. Leukocytosis [C23.550.526]
131. Lithiasis [C23.550.537]
132. Long Term Adverse Effects [C23.550.543]
133. Malacoplakia [C23.550.548]
134. Menstruation Disturbances [C23.550.568]
135. Metaplasia [C23.550.589]
136. Muscle Weakness [C23.550.695]
137. Myotoxicity [C23.550.706]
138. Necrosis [C23.550.717]
139. Neointima [C23.550.722]
140. Neoplastic Processes [C23.550.727]
141. Nerve Degeneration [C23.550.737]
142. Ochronosis [C23.550.744]
143. Ossification, Heterotopic [C23.550.751]
144. Ototoxicity [C23.550.753]
145. Pigmentation Disorders [C23.550.755]
146. Polydipsia [C23.550.759]
147. Postoperative Complications [C23.550.767]
148. Afferent Loop Syndrome [C23.550.767.050]
149. Anastomotic Leak [C23.550.767.071]
150. Breast Cancer Lymphedema [C23.550.767.082]
151. Corneal Endothelial Cell Loss [C23.550.767.093]
152. Coronary-Subclavian Steal Syndrome [C23.550.767.115]
153. Delayed Emergence from Anesthesia [C23.550.767.137]
154. Emergence Delirium [C23.550.767.181]
155. Failed Back Surgery Syndrome [C23.550.767.225]
156. Graft Occlusion, Vascular [C23.550.767.400]
157. Incisional Hernia [C23.550.767.500]
158. Low Anterior Resection Syndrome [C23.550.767.550]
159. Malignant Hyperthermia [C23.550.767.600]
160. Pain, Postoperative [C23.550.767.700]
161. Phantom Limb [C23.550.767.700.500]
162. Postcholecystectomy Syndrome [C23.550.767.775]
163. Postgastrectomy Syndromes [C23.550.767.812]
164. Dumping Syndrome [C23.550.767.812.500]
165. Postoperative Cognitive Complications [C23.550.767.831]
166. Postoperative Hemorrhage [C23.550.767.850]
167. Endoleak [C23.550.767.850.500]
168. Postoperative Nausea and Vomiting [C23.550.767.859]
169. Postpericardiotomy Syndrome [C23.550.767.863]
170. Prosthesis Failure [C23.550.767.865]
171. Implant Capsular Contracture [C23.550.767.865.500]
172. Prosthesis-Related Infections [C23.550.767.868]
173. Reperfusion Injury [C23.550.767.877]
174. Myocardial Reperfusion Injury [C23.550.767.877.500]
175. Post-Cardiac Arrest Syndrome [C23.550.767.877.625]
176. Primary Graft Dysfunction [C23.550.767.877.750]
177. Shock, Surgical [C23.550.767.879]
178. Short Bowel Syndrome [C23.550.767.882]
179. Slit Ventricle Syndrome [C23.550.767.884]
180. Surgical Wound Dehiscence [C23.550.767.887]
181. Surgical Wound Infection [C23.550.767.925]
182. Vasoplegia [C23.550.767.962]

**Intraoperative Complications [C23.550.505]**

66. Blood Loss, Surgical [C23.550.505.300]
67. Intraoperative Awareness [C23.550.505.400]
68. Malignant Hyperthermia [C23.550.505.700]
69. Ischemia [C23.550.513]
70. Chronic Limb-Threatening Ischemia [C23.550.513.178]
71. Infarction [C23.550.513.355]
72. Brain Infarction [C23.550.513.355.250]
73. Hepatic Infarction [C23.550.513.355.500]
74. Myocardial Infarction [C23.550.513.355.750]
75. Pulmonary Infarction [C23.550.513.355.875]
76. Splenic Infarction [C23.550.513.355.937]
77. No-Reflow Phenomenon [C23.550.513.677]
78. Leukoaraiosis [C23.550.522]
79. Leukocytosis [C23.550.526]
80. Lithiasis [C23.550.537]
81. Long-Term Adverse Effects [C23.550.543]
82. Malacoplakia [C23.550.548]
83. Menstruation Disturbances [C23.550.568]
84. Amenorrhea [C23.550.568.500]
85. Dysmenorrhea [C23.550.568.750]
86. Menorrhagia [C23.550.568.875]
87. Oligomenorrhea [C23.550.568.937]
88. Premenstrual Syndrome [C23.550.568.968]
89. Premenstrual Dysphoric Disorder [C23.550.568.968.500]
90. Metaplasia [C23.550.589]
91. Neovascularization, Pathologic [C23.550.589.500]
92. Muscle Weakness [C23.550.695]
93. Dropped Head Syndrome [C23.550.695.500]
94. Myotoxicity [C23.550.706]
95. Necrosis [C23.550.717]
96. Dental Pulp Necrosis [C23.550.717.182]
97. DNA Degradation, Necrotic [C23.550.717.273]
98. Fat Necrosis [C23.550.717.365]
99. Gangrene [C23.550.717.427]
100. Infarction [C23.550.717.489]
101. Brain Infarction [C23.550.717.489.250]
102. Hepatic Infarction [C23.550.717.489.500]
103. Myocardial Infarction [C23.550.717.489.750]
104. Pulmonary Infarction [C23.550.717.489.875]
105. Splenic Infarction [C23.550.717.489.937]
106. Osteonecrosis [C23.550.717.732]
107. Bisphosphonate-Associated Osteonecrosis of the Jaw [C23.550.717.732.183]
108. Femur Head Necrosis [C23.550.717.732.368]
109. Neointima [C23.550.722]
110. Neoplastic Processes [C23.550.727]
111. Anaplasia [C23.550.727.045]
112. Carcinogenesis [C23.550.727.098]
113. Neoplasm Invasiveness [C23.550.727.645]
114. Neoplasm Metastasis [C23.550.727.650]
115. Neoplasm Recurrence, Local [C23.550.727.655]
116. Neoplasm Regression, Spontaneous [C23.550.727.670]
117. Neoplasm, Residual [C23.550.727.700]
118. Oncogene Addiction [C23.550.727.850]
119. Nerve Degeneration [C23.550.737]
120. Retrograde Degeneration [C23.550.737.500]
121. Subacute Combined Degeneration [C23.550.737.625]
122. Wallerian Degeneration [C23.550.737.750]
123. Ochronosis [C23.550.744]
124. Ossification, Heterotopic [C23.550.751]
125. Ossification of Posterior Longitudinal Ligament [C23.550.751.500]
126. Ototoxicity [C23.550.753]
127. Pigmentation Disorders [C23.550.755]
128. Cronkhite-Canada Syndrome [C23.550.755.500]
129. Polydipsia [C23.550.759]
130. Polydipsia, Psychogenic [C23.550.759.500]

**C category Infections** [C01]:

274. Aneurysm, Infected [C01.069]
275. Arthritis, Infectious [C01.100]
276. Asymptomatic Infections [C01.125]
277. Bacterial Infections and Mycoses [C01.150]
278. Bone Diseases, Infectious [C01.160]
279. Breakthrough Infections [C01.175]
280. Cardiovascular Infections [C01.190]
281. Catheter-Related Infections [C01.195]
282. Central Nervous System Infections [C01.207]
283. Coinfection [C01.218]
284. Communicable Diseases [C01.221]
285. Community-Acquired Infections [C01.234]
286. Cross Infection [C01.248]
287. Eye Infections [C01.375]
288. Focal Infection [C01.392]
289. Gingivitis [C01.408]
290. Hepatitis, Animal [C01.436]
291. Intraabdominal Infections [C01.463]
292. Laboratory Infection [C01.503]
293. Latent Infection [C01.550]
294. Opportunistic Infections [C01.597]
295. Parasitic Diseases [C01.610]
296. Pelvic Infection [C01.635]
297. Persistent Infection [C01.645]
298. Poult Enteritis Mortality Syndrome [C01.655]
299. Pregnancy Complications, Infectious [C01.674]
300. Prosthesis-Related Infections [C01.685]
301. Reproductive Tract Infections [C01.730]
302. Respiratory Tract Infections [C01.748]
303. Sepsis [C01.757]
304. Sexually Transmitted Diseases [C01.778]
305. Skin Diseases, Infectious [C01.800]
306. Soft Tissue Infections [C01.820]
307. Suppuration [C01.830]
308. Toxemia [C01.861]
309. Urinary Tract Infections [C01.915]
310. Vaccine-Preventable Diseases [C01.918]
311. Vector Borne Diseases [C01.920]
312. Virus Diseases [C01.925]
313. Waterborne Diseases [C01.936]
314. Wound Infection [C01.947]
315. Zoonoses [C01.973]
316. Neoplasms [C04]
317. Cysts [C04.182]
318. Hamartoma [C04.445]
319. Neoplasms by Histologic Type [C04.557]
320. Neoplasms by Site [C04.588]
321. Neoplasms, Experimental [C04.619]
322. Neoplasms, Hormone-Dependent [C04.626]
323. Neoplasms, Multiple Primary [C04.651]
324. Neoplasms, Post-Traumatic [C04.666]
325. Neoplasms, Radiation-Induced [C04.682]
326. Neoplasms, Second Primary [C04.692]
327. Neoplastic Processes [C04.697]
328. Neoplastic Syndromes, Hereditary [C04.700]
329. Paraneoplastic Syndromes [C04.730]
330. Precancerous Conditions [C04.834]
331. Pregnancy Complications, Neoplastic [C04.850]
332. Musculoskeletal Diseases [C05]
333. Bone Diseases [C05.116]
334. Cartilage Diseases [C05.182]
335. Fasciitis [C05.321]
336. Foot Deformities [C05.330]
337. Foot Diseases [C05.360]
338. Hand Deformities [C05.390]
339. Jaw Diseases [C05.500]
340. Joint Diseases [C05.550]
341. Muscular Diseases [C05.651]
342. Musculoskeletal Abnormalities [C05.660]
343. Rheumatic Diseases [C05.799]
344. Digestive System Diseases [C06]
345. Biliary Tract Diseases [C06.130]
346. Digestive System Abnormalities [C06.198]
347. Digestive System Fistula [C06.267]
348. Digestive System Neoplasms [C06.301]
349. Gastrointestinal Diseases [C06.405]
350. Liver Diseases [C06.552]
351. Pancreatic Diseases [C06.689]
352. Peritoneal Diseases [C06.844]
353. Stomatognathic Diseases [C07]
354. Ankyloglossia [C07.160]
355. Jaw Diseases [C07.320]
356. Mouth Diseases [C07.465]
357. Pharyngeal Diseases [C07.550]
358. Stomatognathic System Abnormalities [C07.650]
359. Temporomandibular Joint Disorders [C07.678]
360. Tooth Diseases [C07.793]
361. Respiratory Tract Diseases [C08]
362. Bronchial Diseases [C08.127]
363. Ciliary Motility Disorders [C08.200]
364. Granuloma, Respiratory Tract [C08.280]
365. Laryngeal Diseases [C08.360]
366. Lung Diseases [C08.381]
367. Nose Diseases [C08.460]
368. Pleural Diseases [C08.528]
369. Respiration Disorders [C08.618]
370. Respiratory Hypersensitivity [C08.674]
371. Respiratory System Abnormalities [C08.695]
372. Respiratory Tract Fistula [C08.702]
373. Respiratory Tract Infections [C08.730]
374. Respiratory Tract Neoplasms [C08.785]
375. Thoracic Diseases [C08.846]
376. Tracheal Diseases [C08.907]
377. Otorhinolaryngologic Diseases [C09]
378. Ciliary Motility Disorders [C09.150]
379. Ear Diseases [C09.218]
380. Laryngeal Diseases [C09.400]
381. Nose Diseases [C09.603]
382. Otorhinolaryngologic Neoplasms [C09.647]
383. Pharyngeal Diseases [C09.775]
384. Nervous System Diseases [C10]
385. Autoimmune Diseases of the Nervous System [C10.114]
386. Autonomic Nervous System Diseases [C10.177]
387. Central Nervous System Diseases [C10.228]
388. Chronobiology Disorders [C10.281]
389. Cranial Nerve Diseases [C10.292]
390. Demyelinating Diseases [C10.314]
391. Nervous System Malformations [C10.500]
392. Nervous System Neoplasms [C10.551]
393. Neurocutaneous Syndromes [C10.562]
394. Neurodegenerative Diseases [C10.574]
395. Neuroinflammatory Diseases [C10.586]
396. Neurologic Manifestations [C10.597]
397. Neuromuscular Diseases [C10.668]
398. Neurotoxicity Syndromes [C10.720]
399. Restless Legs Syndrome [C10.803]
400. Sleep Wake Disorders [C10.886]
401. Trauma, Nervous System [C10.900]
402. Eye Diseases [C11]
403. Asthenopia [C11.093]
404. Cogan Syndrome [C11.180]
405. Conjunctival Diseases [C11.187]
406. Corneal Diseases [C11.204]
407. Eye Abnormalities [C11.250]
408. Eye Diseases, Hereditary [C11.270]
409. Eye Hemorrhage [C11.290]
410. Eye Infections [C11.294]
411. Eye Injuries [C11.297]
412. Eye Manifestations [C11.300]
413. Eye Neoplasms [C11.319]
414. Eyelid Diseases [C11.338]
415. Lacrimal Apparatus Diseases [C11.496]
416. Lens Diseases [C11.510]
417. Ocular Hypertension [C11.525]
418. Ocular Hypotension [C11.540]
419. Ocular Motility Disorders [C11.590]
420. Optic Nerve Diseases [C11.640]
421. Orbital Diseases [C11.675]
422. Pupil Disorders [C11.710]
423. Refractive Errors [C11.744]
424. Retinal Diseases [C11.768]
425. Scleral Diseases [C11.790]
426. Uveal Diseases [C11.941]
427. Vision Disorders [C11.966]
428. Vitreous Detachment [C11.980]
429. Urogenital Diseases [C12]
430. Female Urogenital Diseases and Pregnancy Complications [C12.050]
431. Genital Diseases [C12.100]
432. Male Urogenital Diseases [C12.200]
433. Pelvic Floor Disorders [C12.400]
434. Tuberculosis, Urogenital [C12.600]
435. Urogenital Abnormalities [C12.800]
436. Urogenital Neoplasms [C12.900]
437. Urologic Diseases [C12.950]
438. Cardiovascular Diseases [C14]
439. Cardiovascular Abnormalities [C14.240]
440. Cardiovascular Infections [C14.260]
441. Heart Diseases [C14.280]
442. Pregnancy Complications, Cardiovascular [C14.583]
443. Vascular Diseases [C14.907]
444. Hemic and Lymphatic Diseases [C15]
445. Hematologic Diseases [C15.378]
446. Lymphatic Diseases [C15.604]
447. Congenital, Hereditary, and Neonatal Diseases and Abnormalities [C16]
448. Congenital Abnormalities [C16.131]
449. Fetal Diseases [C16.300]
450. Genetic Diseases, Inborn [C16.320]
451. Infant, Newborn, Diseases [C16.614]
452. Skin and Connective Tissue Diseases [C17]
453. Connective Tissue Diseases [C17.300]
454. Skin Diseases [C17.800]
455. Nutritional and Metabolic Diseases [C18]
456. Metabolic Diseases [C18.452]
457. Nutrition Disorders [C18.654]
458. Endocrine System Diseases [C19]
459. Adrenal Gland Diseases [C19.053]
460. Bone Diseases, Endocrine [C19.149]
461. Diabetes Mellitus [C19.246]
462. Dwarfism [C19.297]
463. Endocrine Gland Neoplasms [C19.344]
464. Gonadal Disorders [C19.391]
465. Parathyroid Diseases [C19.642]
466. Pituitary Diseases [C19.700]
467. Polyendocrinopathies, Autoimmune [C19.787]
468. Thyroid Diseases [C19.874]
469. Tuberculosis, Endocrine [C19.927]
470. Immune System Diseases [C20]
471. Autoimmune Diseases [C20.111]
472. Erythroblastosis, Fetal [C20.306]
473. Glomerulonephritis, Membranoproliferative [C20.425]
474. Graft vs Host Disease [C20.452]
475. Hypersensitivity [C20.543]
476. Immune Reconstitution Inflammatory Syndrome [C20.608]
477. Immunologic Deficiency Syndromes [C20.673]
478. Immunoproliferative Disorders [C20.683]
479. Mast Cell Activation Disorders [C20.762]
480. Purpura, Thrombocytopenic [C20.841]
481. Transfusion Reaction [C20.920]
482. Disorders of Environmental Origin [C21]
483. Environmental Illness [C21.223]
484. Preconception Injuries [C21.676]
485. Animal Diseases [C22]
486. Pathological Conditions, Signs and Symptoms [C23]
487. Morphological and Microscopic Findings [C23.149]
488. Pathological Conditions, Anatomical [C23.300]
489. Pathologic Processes [C23.550]
490. Signs and Symptoms [C23.888]
491. Occupational Diseases [C24]
492. Chemically-Induced Disorders [C25]
493. Wounds and Injuries [C26]
494. Abdominal Injuries [C26.017]
495. Accidental Injuries [C26.040]
496. Amputation, Traumatic [C26.062]
497. Arm Injuries [C26.088]
498. Asphyxia [C26.103]
499. Athletic Injuries [C26.115]
500. Back Injuries [C26.117]
501. Barotrauma [C26.120]
502. Birth Injuries [C26.141]
503. Bites and Stings [C26.176]
504. Burns [C26.200]
505. Cold Injury [C26.212]
506. Contrecoup Injury [C26.224]
507. Crush Injuries [C26.257]
508. Joint Dislocations [C26.289]
509. Drowning [C26.304]
510. Electric Injuries [C26.324]
511. Esophageal Perforation [C26.348]
512. Extravasation of Diagnostic and Therapeutic Materials [C26.371]
513. Foreign Bodies [C26.392]
514. Fractures, Bone [C26.404]
515. Fractures, Cartilage [C26.411]
516. Frostbite [C26.417]
517. Hand Injuries [C26.448]
518. Heat Stress Disorders [C26.522]
519. Hip Injuries [C26.531]
520. Lacerations [C26.540]
521. Leg Injuries [C26.558]
522. Microtrauma, Physical [C26.599]
523. Multiple Trauma [C26.640]
524. Nasal Septal Perforation [C26.670]
525. Neck Injuries [C26.700]
526. Occupational Injuries [C26.716]
527. Radiation Injuries [C26.733]
528. Reinjuries [C26.741]
529. Retropneumoperitoneum [C26.748]
530. Rupture [C26.761]
531. Self Mutilation [C26.780]
532. Shock, Traumatic [C26.797]
533. Shoulder Injuries [C26.803]
534. Soft Tissue Injuries [C26.808]
535. Spinal Cord Injuries [C26.819]
536. Sprains and Strains [C26.844]
537. Surgical Wound [C26.859]
538. Tendon Injuries [C26.874]
539. Thoracic Injuries [C26.891]
540. Tooth Injuries [C26.900]
541. Trauma, Nervous System [C26.915]
542. Tympanic Membrane Perforation [C26.930]
543. Vascular System Injuries [C26.940]
544. War-Related Injuries [C26.946]
545. Wounds, Nonpenetrating [C26.974]
546. Wounds, Penetrating [C26.986]

\###

**Output example**:

[

{

"VirtualPatientID": "VP1",

"PredictedComplications": [

{"ComplicationName": "Surgical Wound Infection", "MeSHTreeNumber": "C23.550.767.925", "NormalizedProbability": 15},

{"ComplicationName": "Postoperative Hemorrhage", "MeSHTreeNumber": "C23.550.767.850", "NormalizedProbability": 12},

{"ComplicationName": "Abdominal Abscess", "MeSHTreeNumber": "C01.830.025.020", "NormalizedProbability": 11},

{"ComplicationName": "Biliary Fistula", "MeSHTreeNumber": "C06.267.500", "NormalizedProbability": 10},

{"ComplicationName": "Acute Pancreatitis", "MeSHTreeNumber": "C06.689.150", "NormalizedProbability": 10},

{"ComplicationName": "Sepsis", "MeSHTreeNumber": "C01.757", "NormalizedProbability": 9},

{"ComplicationName": "Shock, Surgical", "MeSHTreeNumber": "C23.550.767.879", "NormalizedProbability": 9},

{"ComplicationName": "Postoperative Nausea and Vomiting", "MeSHTreeNumber": "C23.550.767.859", "NormalizedProbability": 8},

{"ComplicationName": "Incisional Hernia", "MeSHTreeNumber": "C23.550.767.500", "NormalizedProbability": 8},

{"ComplicationName": "Pain, Postoperative", "MeSHTreeNumber": "C23.550.767.700", "NormalizedProbability": 8}

]

},

{

"VirtualPatientID": "VP2",

"PredictedComplications": [

{"ComplicationName": "Postoperative Hemorrhage", "MeSHTreeNumber": "C23.550.767.850", "NormalizedProbability": 14},

{"ComplicationName": "Surgical Wound Infection", "MeSHTreeNumber": "C23.550.767.925", "NormalizedProbability": 13},

{"ComplicationName": "Abdominal Abscess", "MeSHTreeNumber": "C01.830.025.020", "NormalizedProbability": 12},

{"ComplicationName": "Biliary Fistula", "MeSHTreeNumber": "C06.267.500", "NormalizedProbability": 11},

{"ComplicationName": "Acute Pancreatitis", "MeSHTreeNumber": "C06.689.150", "NormalizedProbability": 10},

{"ComplicationName": "Sepsis", "MeSHTreeNumber": "C01.757", "NormalizedProbability": 10},

{"ComplicationName": "Shock, Surgical", "MeSHTreeNumber": "C23.550.767.879", "NormalizedProbability": 8},

{"ComplicationName": "Postoperative Nausea and Vomiting", "MeSHTreeNumber": "C23.550.767.859", "NormalizedProbability": 7},

{"ComplicationName": "Incisional Hernia", "MeSHTreeNumber": "C23.550.767.500", "NormalizedProbability": 5},

{"ComplicationName": "Pain, Postoperative", "MeSHTreeNumber": "C23.550.767.700", "NormalizedProbability": 5}

]

},

{

"VirtualPatientID": "VP3",

"PredictedComplications": [

{"ComplicationName": "Abdominal Abscess", "MeSHTreeNumber": "C01.830.025.020", "NormalizedProbability": 16},

{"ComplicationName": "Surgical Wound Infection", "MeSHTreeNumber": "C23.550.767.925", "NormalizedProbability": 14},

{"ComplicationName": "Postoperative Hemorrhage", "MeSHTreeNumber": "C23.550.767.850", "NormalizedProbability": 12},

{"ComplicationName": "Biliary Fistula", "MeSHTreeNumber": "C06.267.500", "NormalizedProbability": 11},

{"ComplicationName": "Acute Pancreatitis", "MeSHTreeNumber": "C06.689.150", "NormalizedProbability": 10},

{"ComplicationName": "Sepsis", "MeSHTreeNumber": "C01.757", "NormalizedProbability": 9},

{"ComplicationName": "Shock, Surgical", "MeSHTreeNumber": "C23.550.767.879", "NormalizedProbability": 8},

{"ComplicationName": "Postoperative Nausea and Vomiting", "MeSHTreeNumber": "C23.550.767.859", "NormalizedProbability": 6},

{"ComplicationName": "Pain, Postoperative", "MeSHTreeNumber": "C23.550.767.700", "NormalizedProbability": 4},

{"ComplicationName": "Incisional Hernia", "MeSHTreeNumber": "C23.550.767.500", "NormalizedProbability": 4}

]

},

{

"VirtualPatientID": "VP4",

"PredictedComplications": [

{"ComplicationName": "Surgical Wound Infection", "MeSHTreeNumber": "C23.550.767.925", "NormalizedProbability": 17},

{"ComplicationName": "Postoperative Hemorrhage", "MeSHTreeNumber": "C23.550.767.850", "NormalizedProbability": 14},

{"ComplicationName": "Biliary Fistula", "MeSHTreeNumber": "C06.267.500", "NormalizedProbability": 12},

{"ComplicationName": "Abdominal Abscess", "MeSHTreeNumber": "C01.830.025.020", "NormalizedProbability": 11},

{"ComplicationName": "Acute Pancreatitis", "MeSHTreeNumber": "C06.689.150", "NormalizedProbability": 10},

{"ComplicationName": "Sepsis", "MeSHTreeNumber": "C01.757", "NormalizedProbability": 8},

{"ComplicationName": "Shock, Surgical", "MeSHTreeNumber": "C23.550.767.879", "NormalizedProbability": 7},

{"ComplicationName": "Postoperative Nausea and Vomiting", "MeSHTreeNumber": "C23.550.767.859", "NormalizedProbability": 6},

{"ComplicationName": "Pain, Postoperative", "MeSHTreeNumber": "C23.550.767.700", "NormalizedProbability": 5},

{"ComplicationName": "Incisional Hernia", "MeSHTreeNumber": "C23.550.767.500", "NormalizedProbability": 5}

]

},

{

"VirtualPatientID": "VP5",

"PredictedComplications": [

{"ComplicationName": "Abdominal Abscess", "MeSHTreeNumber": "C01.830.025.020", "NormalizedProbability": 16},

{"ComplicationName": "Surgical Wound Infection", "MeSHTreeNumber": "C23.550.767.925", "NormalizedProbability": 14},

{"ComplicationName": "Postoperative Hemorrhage", "MeSHTreeNumber": "C23.550.767.850", "NormalizedProbability": 12},

{"ComplicationName": "Biliary Fistula", "MeSHTreeNumber": "C06.267.500", "NormalizedProbability": 11},

{"ComplicationName": "Acute Pancreatitis", "MeSHTreeNumber": "C06.689.150", "NormalizedProbability": 10},

{"ComplicationName": "Sepsis", "MeSHTreeNumber": "C01.757", "NormalizedProbability": 9},

{"ComplicationName": "Shock, Surgical", "MeSHTreeNumber": "C23.550.767.879", "NormalizedProbability": 8},

{"ComplicationName": "Postoperative Nausea and Vomiting", "MeSHTreeNumber": "C23.550.767.859", "NormalizedProbability": 5},

{"ComplicationName": "Pain, Postoperative", "MeSHTreeNumber": "C23.550.767.700", "NormalizedProbability": 4},

{"ComplicationName": "Incisional Hernia", "MeSHTreeNumber": "C23.550.767.500", "NormalizedProbability": 4}

]

},

{

"VirtualPatientID": "VP6",

"PredictedComplications": [

{"ComplicationName": "Surgical Wound Infection", "MeSHTreeNumber": "C23.550.767.925", "NormalizedProbability": 16},

{"ComplicationName": "Abdominal Abscess", "MeSHTreeNumber": "C01.830.025.020", "NormalizedProbability": 15},

{"ComplicationName": "Postoperative Hemorrhage", "MeSHTreeNumber": "C23.550.767.850", "NormalizedProbability": 12},

{"ComplicationName": "Biliary Fistula", "MeSHTreeNumber": "C06.267.500", "NormalizedProbability": 11},

{"ComplicationName": "Acute Pancreatitis", "MeSHTreeNumber": "C06.689.150", "NormalizedProbability": 10},

{"ComplicationName": "Sepsis", "MeSHTreeNumber": "C01.757", "NormalizedProbability": 8},

{"ComplicationName": "Shock, Surgical", "MeSHTreeNumber": "C23.550.767.879", "NormalizedProbability": 7},

{"ComplicationName": "Postoperative Nausea and Vomiting", "MeSHTreeNumber": "C23.550.767.859", "NormalizedProbability": 6},

{"ComplicationName": "Pain, Postoperative", "MeSHTreeNumber": "C23.550.767.700", "NormalizedProbability": 5},

{"ComplicationName": "Incisional Hernia", "MeSHTreeNumber": "C23.550.767.500", "NormalizedProbability": 4}

]

},

{

"VirtualPatientID": "VP7",

"PredictedComplications": [

{"ComplicationName": "Abdominal Abscess", "MeSHTreeNumber": "C01.830.025.020", "NormalizedProbability": 17},

{"ComplicationName": "Postoperative Hemorrhage", "MeSHTreeNumber": "C23.550.767.850", "NormalizedProbability": 14},

{"ComplicationName": "Surgical Wound Infection", "MeSHTreeNumber": "C23.550.767.925", "NormalizedProbability": 13},

{"ComplicationName": "Biliary Fistula", "MeSHTreeNumber": "C06.267.500", "NormalizedProbability": 11},

{"ComplicationName": "Acute Pancreatitis", "MeSHTreeNumber": "C06.689.150", "NormalizedProbability": 10},

{"ComplicationName": "Sepsis", "MeSHTreeNumber": "C01.757", "NormalizedProbability": 9},

{"ComplicationName": "Shock, Surgical", "MeSHTreeNumber": "C23.550.767.879", "NormalizedProbability": 8},

{"ComplicationName": "Postoperative Nausea and Vomiting", "MeSHTreeNumber": "C23.550.767.859", "NormalizedProbability": 5},

{"ComplicationName": "Pain, Postoperative", "MeSHTreeNumber": "C23.550.767.700", "NormalizedProbability": 5},

{"ComplicationName": "Incisional Hernia", "MeSHTreeNumber": "C23.550.767.500", "NormalizedProbability": 4}

]

},

{

"VirtualPatientID": "VP8",

"PredictedComplications": [

{"ComplicationName": "Surgical Wound Infection", "MeSHTreeNumber": "C23.550.767.925", "NormalizedProbability": 16},

{"ComplicationName": "Postoperative Hemorrhage", "MeSHTreeNumber": "C23.550.767.850", "NormalizedProbability": 14},

{"ComplicationName": "Abdominal Abscess", "MeSHTreeNumber": "C01.830.025.020", "NormalizedProbability": 12},

{"ComplicationName": "Biliary Fistula", "MeSHTreeNumber": "C06.267.500", "NormalizedProbability": 11},

{"ComplicationName": "Acute Pancreatitis", "MeSHTreeNumber": "C06.689.150", "NormalizedProbability": 10},

{"ComplicationName": "Sepsis", "MeSHTreeNumber": "C01.757", "NormalizedProbability": 9},

{"ComplicationName": "Shock, Surgical", "MeSHTreeNumber": "C23.550.767.879", "NormalizedProbability": 8},

{"ComplicationName": "Postoperative Nausea and Vomiting", "MeSHTreeNumber": "C23.550.767.859", "NormalizedProbability": 5},

{"ComplicationName": "Pain, Postoperative", "MeSHTreeNumber": "C23.550.767.700", "NormalizedProbability": 5},

{"ComplicationName": "Incisional Hernia", "MeSHTreeNumber": "C23.550.767.500", "NormalizedProbability": 5}

]

},

{

"VirtualPatientID": "VP9",

"PredictedComplications": [

{"ComplicationName": "Abdominal Abscess", "MeSHTreeNumber": "C01.830.025.020", "NormalizedProbability": 17},

{"ComplicationName": "Surgical Wound Infection", "MeSHTreeNumber": "C23.550.767.925", "NormalizedProbability": 15},

{"ComplicationName": "Postoperative Hemorrhage", "MeSHTreeNumber": "C23.550.767.850", "NormalizedProbability": 12},

{"ComplicationName": "Biliary Fistula", "MeSHTreeNumber": "C06.267.500", "NormalizedProbability": 10},

{"ComplicationName": "Acute Pancreatitis", "MeSHTreeNumber": "C06.689.150", "NormalizedProbability": 10},

{"ComplicationName": "Sepsis", "MeSHTreeNumber": "C01.757", "NormalizedProbability": 8},

{"ComplicationName": "Shock, Surgical", "MeSHTreeNumber": "C23.550.767.879", "NormalizedProbability": 7},

{"ComplicationName": "Postoperative Nausea and Vomiting", "MeSHTreeNumber": "C23.550.767.859", "NormalizedProbability": 6},

{"ComplicationName": "Pain, Postoperative", "MeSHTreeNumber": "C23.550.767.700", "NormalizedProbability": 5},

{"ComplicationName": "Incisional Hernia", "MeSHTreeNumber": "C23.550.767.500", "NormalizedProbability": 5}

]

},

{

"VirtualPatientID": "VP10",

"PredictedComplications": [

{"ComplicationName": "Surgical Wound Infection", "MeSHTreeNumber": "C23.550.767.925", "NormalizedProbability": 16},

{"ComplicationName": "Abdominal Abscess", "MeSHTreeNumber": "C01.830.025.020", "NormalizedProbability": 15},

{"ComplicationName": "Postoperative Hemorrhage", "MeSHTreeNumber": "C23.550.767.850", "NormalizedProbability": 12},

{"ComplicationName": "Biliary Fistula", "MeSHTreeNumber": "C06.267.500", "NormalizedProbability": 11},

{"ComplicationName": "Acute Pancreatitis", "MeSHTreeNumber": "C06.689.150", "NormalizedProbability": 10},

{"ComplicationName": "Sepsis", "MeSHTreeNumber": "C01.757", "NormalizedProbability": 8},

{"ComplicationName": "Shock, Surgical", "MeSHTreeNumber": "C23.550.767.879", "NormalizedProbability": 7},

{"ComplicationName": "Postoperative Nausea and Vomiting", "MeSHTreeNumber": "C23.550.767.859", "NormalizedProbability": 6},

{"ComplicationName": "Pain, Postoperative", "MeSHTreeNumber": "C23.550.767.700", "NormalizedProbability": 5},

{"ComplicationName": "Incisional Hernia", "MeSHTreeNumber": "C23.550.767.500", "NormalizedProbability": 5}

]

}

]

Prompt 4C

**Summarize Complications Across Virtual Patients**

**Input:** JSON array of virtual patients. Each virtual patient contains a list of predicted complications in the following format:

[

{

"VirtualPatientID": "VP1",

"PredictedComplications": [

{"ComplicationName": "...", "MeSHTreeNumber": "...", "NormalizedProbability": ...},

...

]

},

...

]

**##**

**The JSON array of virtual patients is: [HERE]**

**##**

**Task:**

1. For each unique complication across all virtual patients, calculate:
   - VPCount: Number of virtual patients for which the complication was predicted.
   - AverageNormalizedProbability: Average of the NormalizedProbability across all virtual patients where it was predicted.
   - MeSHTreeNumber: The corresponding MeSH tree number for the complication.
2. After calculating the averages, rescale the AverageNormalizedProbability values so that the sum across all complications equals 100.
3. Return a JSON array of dictionaries with the following fields:
   - ComplicationName
   - MeSHTreeNumber
   - VPCount
   - AverageNormalizedProbability (after rescaling so the sum equals 100)

**Output format (JSON):**

[

{

"ComplicationName": "Complication A",

"MeSHTreeNumber": "C23.550.767.850",

"VPCount": 3,

"AverageNormalizedProbability": 15.2

},

...

]

**Additional Instructions:**

- Include the MeSH tree number exactly as it appears in the input.
- Sort the output by VPCount in descending order, then by AverageNormalizedProbability.
- Only include complications that appear in at least one virtual patient.
- Ensure that the sum of all AverageNormalizedProbability values equals 100.

\###

Output example:

[

{

"ComplicationName": "Surgical Wound Infection",

"MeSHTreeNumber": "C23.550.767.925",

"VPCount": 10,

"AverageNormalizedProbability": 15.5

},

{

"ComplicationName": "Abdominal Abscess",

"MeSHTreeNumber": "C01.830.025.020",

"VPCount": 10,

"AverageNormalizedProbability": 14.8

},

{

"ComplicationName": "Postoperative Hemorrhage",

"MeSHTreeNumber": "C23.550.767.850",

"VPCount": 10,

"AverageNormalizedProbability": 12.2

},

{

"ComplicationName": "Biliary Fistula",

"MeSHTreeNumber": "C06.267.500",

"VPCount": 10,

"AverageNormalizedProbability": 10.7

},

{

"ComplicationName": "Acute Pancreatitis",

"MeSHTreeNumber": "C06.689.150",

"VPCount": 10,

"AverageNormalizedProbability": 10.2

},

{

"ComplicationName": "Sepsis",

"MeSHTreeNumber": "C01.757",

"VPCount": 10,

"AverageNormalizedProbability": 8.5

},

{

"ComplicationName": "Shock, Surgical",

"MeSHTreeNumber": "C23.550.767.879",

"VPCount": 10,

"AverageNormalizedProbability": 7.8

},

{

"ComplicationName": "Postoperative Nausea and Vomiting",

"MeSHTreeNumber": "C23.550.767.859",

"VPCount": 10,

"AverageNormalizedProbability": 5.6

},

{

"ComplicationName": "Pain, Postoperative",

"MeSHTreeNumber": "C23.550.767.700",

"VPCount": 10,

"AverageNormalizedProbability": 4.8

},

{

"ComplicationN

**Prompt 5.**

The agent uses this prompt to predict the the most likely complication of a given surgical procedure.

**Instructions:**

Below is a list of surgical procedures.

For each surgical procedure, name the most likely complication and the least likely complication

Present each complication in **short, standard clinical language** and, when available, link it to the **MeSH descriptor** (Medical Subject Headings, NLM controlled vocabulary).

Output format as in the example

**##**

**Example:**

**Procedure: Tracheostomy**

**Most likely complication**

- **Clinical term:** **Tracheostomy Tube Obstruction**
- **Definition (short clinical language):** Blockage of the artificial airway due to a mucus plug, crusting, or kinking of the tracheostomy tube.
- **MeSH Descriptor:**
  - *Tracheostomy/adverse effects [D014141]*
  - *Airway Obstruction [D000210]*

**Least likely complication**

- **Clinical term:** **Tracheoesophageal Fistula**
- **Definition (short clinical language):** An abnormal communication between the trachea and esophagus, usually arising as a rare late complication of tracheostomy or prolonged intubation.
- **MeSH Descriptor:**
  - *Tracheoesophageal Fistula [D014143]*

**Prompt 6**

**Classifying surgical complications**

**Instructions**

You are given two surgical complications:

Complication 1: [Observed Complication Name HERE]

Complication 2: [Predicted Complication Name HERE]

Use the “List of Surgical Complication Categories”

Classify Complications 1 and 2 internally

If they are classified into the same category, return “1”; otherwise, return “0”.

**##**

**Example:**

Complication 1: Postoperative wound infection

Complication 2: Surgical site infection (SSI)

Output: “1” (Category: Infectious)

**###**

**List of Surgical Complication Categories**

**1. Infectious**

**Definition:** Complications caused by microbial contamination or infection.
**Examples:** Surgical site infection, intra-abdominal abscess, peritonitis

**2. Hemorrhagic / Hematologic**

**Definition:** Complications related to bleeding, hematoma, or failure of hemostasis.
**Examples:** Postoperative hemorrhage, hematoma

**3. Mechanical / Obstructive / Technical Injury**

**Definition:** Complications caused by retained material, physical blockage, device malfunction, or intraoperative injury.
**Examples:** Retained stone, hernia, bowel obstruction, anastomotic stricture

**4. Fistula / Leak**

**Definition:** Abnormal connection or leakage from a hollow viscus or surgical anastomosis.
**Examples:** Bile leak, pancreatic fistula, anastomotic leak

**5. Functional / Motility-related**

**Definition:** Complications characterized by failure of normal organ function despite intact anatomical structure.
**Examples:** Postoperative ileus, urinary retention

**6. Thrombotic / Vascular**

**Definition:** Complications due to vascular compromise, occlusion, or clotting disorders.
**Examples:** Deep vein thrombosis (DVT), portal vein thrombosis, embolism

**7. Anesthetic / Medication-related**

**Definition:** Complications arising from anesthesia, airway management, or perioperative medications.
**Examples:** Malignant hyperthermia, airway injury, drug reactions

**8. Neurological**

**Definition:** Complications involving injury to nerves or central nervous system structures.
**Examples:** Peripheral nerve palsy, stroke, spinal cord injury

**9. Cardiopulmonary**

**Definition:** Complications of the heart or lungs occurring perioperatively or postoperatively.
**Examples:** Myocardial infarction, pulmonary embolism, pneumonia

**10. Wound-related (non-infectious)**

**Definition:** Complications of wound healing not caused by infection.
**Examples:** Dehiscence, seroma, hypertrophic scar

**11. Metabolic / Endocrine / Electrolyte**

**Definition:** Complications due to metabolic imbalance, endocrine dysfunction, or electrolyte disturbances.
**Examples:** Hypocalcemia (post-thyroidectomy), adrenal insufficiency, hyponatremia, hyperglycemia

**12. Oncologic Recurrence / Progression**

**Definition:** Recurrence or persistence of malignant disease following surgical treatment.
**Examples:** Local tumor recurrence, recurrent hepatic neoplasm

**13. Pain & Rehabilitation-related**

**Definition:** Persistent or unexpected pain and impaired functional recovery after surgery.
**Examples:** Chronic post-surgical pain, neuropathic pain, phantom limb pain

| **Age Range** | **Age Group** |
| --- | --- |
| **<18** | 1 - Pediatric |
| **18–39** | 2 - Young Adult |
| **40–59** | 3 - Mid-age Adult |
| **60–69** | 4 - Older Adult |
| **70–79** | 5 - Elderly |
| **80+** | 6 - Very Elderly |

Table 1. Age groups

BIBAS UI version

This prompt can be directly inserted into the UI of any LLM. It does not really run the simulation. However, it prompts the LLM to think *as if* it runs a simulation. This prompt gained 27% HIT rate.

To apply the prompt, the input is:

1. The PATIENT’S DESCRIPTION
2. The PATIENT’S DIAGNOSIS
3. The PLANNED SURGERY
4. TIMING of expected complication (IMMEDIATE OR EARLY vs. LATE OR LONG-TERM)

First, you run the RISK FACTORS ANALYSIS prompt:

**Identifying Risk Factors that might Lead to Surgical Complications**

**Definition:**
A *risk factor* is any patient-related, disease-related, anatomical, physiological, or procedural characteristic present *before* the surgical intervention that increases the likelihood of **intraoperative or postoperative surgical complications** (NOT general adverse outcomes).

**Task:** You are an expert in clinical risk analysis. Given a PATIENT’S DESCRIPTION, the patient’s DIAGNOSIS, and a PLANNED SURGERY, your task is to identify and extract all **clinically relevant preoperative risk factors** that have **established, strong correlations** with surgical complications from that intervention.

**Input:**

- PATIENT’S DESCRIPTION: {PATIENT’S DESCRIPTION HERE}
- DIAGNOSIS: {DIAGNOSIS HERE}
- PLANNED SURGERY: {PLANNED SURGERY HERE}

**Requirements:**

6. Focus specifically on risk factors for **surgical complications,** including:
   - Intraoperative complications (bleeding, organ injury, conversion to open surgery, anesthetic complications)
   - Postoperative complications (wound infection, bile leaks, bleeding, pneumonia, thromboembolism, delayed recovery)
   - NOT general medical outcomes or mortality risk
7. Only include risk factors with **established, strong clinical evidence** for increasing surgical complication risk:
   - Must have documented correlation in surgical literature
   - Exclude weak, questionable, or indirect associations
   - Exclude factors that primarily affect general medical outcomes rather than surgical complications
8. Extract relevant information, including:
   - Patient demographics (age, BMI if available) - only if strongly correlated
   - Comorbidities with established surgical risk (e.g., active infection, severe cardiac disease, coagulopathy)
   - Anatomical and pathological conditions affecting surgical difficulty (e.g., active inflammation, anatomical distortion)
   - Laboratory abnormalities directly affecting surgery (e.g., coagulation defects, severe organ dysfunction)
   - Imaging findings indicating surgical complexity
   - Clinical status indicators affecting surgical risk (e.g., hemodynamic instability, active infection)
9. Classify each risk factor by type:
   - Demographic, Comorbidity, Anatomical/Pathological, Laboratory/Physiological, Procedural history, Clinical status
10. Perform internal quality control to ensure extracted risk factors:
    - Have a **strong, established** correlation with surgical complications
    - Are specific to the surgical intervention being performed
    - Directly impact intraoperative or postoperative surgical outcomes

**Output Format:** Provide only the final list of extracted risk factors in this exact format: [Risk Factor ICD-10-CM Name, Risk Factor Type]

**Notes:**

- Include only risk factors with robust clinical evidence for surgical complication correlation
- Exclude factors with weak, questionable, or primarily non-surgical outcome associations
- Provide no additional commentary or explanation beyond the required output format

**Example:**

**PATIENT’S DESCRIPTION:** {Maria G., a 74-year-old female, presented to the emergency department with a 2-day history of severe epigastric pain radiating to the back, associated with nausea and multiple episodes of non-bloody vomiting. She reported decreased appetite and subjective fever. Her past medical history included gallstones, hyperlipidemia, and coronary artery disease. On physical examination, the patient was febrile (38.7°C), with tenderness in the right upper quadrant and epigastrium. Laboratory findings revealed elevated white blood cell count, increased serum lipase (1120 U/L), and mildly abnormal liver function tests. Abdominal ultrasound showed gallstones without ductal dilatation, while CT imaging confirmed acute gallstone pancreatitis, with signs of peripancreatic fat stranding and inflammation. The patient was admitted and initially managed with intravenous fluids, analgesia, and bowel rest. On hospital day 3, due to persistent symptoms and recurrent pain, the surgical team proceeded with—}

**DIAGNOSIS**: {**Gallstone Pancreatitis**}

**PLANNED SURGERY**: {**laparoscopic cholecystectomy**}

**Output:**

**RISK FACTORS:**

[Age 74 years, Demographic]

[Acute pancreatitis, Anatomical/Pathological]

[Elevated white blood cell count, Laboratory/Physiological]

[Peripancreatic inflammation, Anatomical/Pathological]

***

Use the output of RISK FACTORS and run prompts A to C. These prompts appear on the next page.

Instructions: Run Prompts A to C sequentially and internally.

Generate ONLY the final output of prompt C.

**PROMPT A: Virtual Patients Simulation**

**Introduction:**

- A virtual patient is a representation of a patient’s clinical profile

with the purpose of simulating and predicting health outcomes.

- A **patient’s profile** includes his/her (1) DIAGNOSIS, (2) planned SURGICAL INTERVENTION, and (3) RISK FACTORS.
- Your task is to generate a Virtual Patients Cohort from a patient’s profile.

**The structure of the prompt:**

- **Module 1** determines the data type of each RISK FACTOR.
- **Module 2** identifies the distribution of each RISK FACTOR.
- **Module 3** generates up to 50 unique virtual patients from the patient's profile.

**Your input is:**

(5) The patient's RISK FACTORS: [PATIENT’S risk factors HERE]
(6) The patient’s DIAGNOSIS: [DIAGNOSIS HERE]
(7) The PLANNED SURGERY: [PLANNED SURGERY HERE]
(8) TIMING of the complication: [TIMING HERE]

**Note**: The TIMING of a complication corresponds with the following typology:

**1. Immediate Complications**

- **Timing:** During surgery or within **24 hours**
- **Examples:**
  - Hemorrhage (intraoperative or postoperative bleeding)
  - Anesthesia-related issues (e.g., aspiration, cardiac events)
  - Injury to adjacent structures
  - Acute allergic reactions

**2. Early Complications**

- **Timing:** Within **24 hours to 30 days** after surgery
- **Examples:**
  - Surgical site infection (SSI)
  - Pneumonia
  - Deep vein thrombosis (DVT) or pulmonary embolism (PE)
  - Paralytic ileus or bowel obstruction
  - Urinary leakage or fistula
  - Acute kidney injury
  - Wound dehiscence

**3. Late Complications**

- **Timing:** After **30 days**, often up to **months post-op**
- **Examples:**
  - Anastomotic strictures
  - Hernia at the surgical site
  - Ureteral obstruction or stricture
  - Chronic pain
  - Delayed infections (e.g., deep abscess)

**4. Long-term (or Chronic) Complications**

- **Timing:** **Months to years** after surgery
- **Examples:**
  - **Neoplasm recurrence**
  - Chronic kidney disease (from urinary diversion)
  - Metabolic disturbances (e.g., acidosis with ileal conduit)
  - Sexual dysfunction
  - Long-term urinary incontinence or retention

**Final INTERNAL output for PROMPT A:**

- A list of the virtual patients, each with his/her unique parameters of risk factors.

___________________________________________________________________

**Internal quality check:**

- Perform an internal quality check
- Ensure you generated DIFFERENT and unique virtual patients from each **patient’s profile**.
- Ensure that each module has been appropriately applied

___________________________________________________________________

**THE MODULES**

**Module 1. Identify the Data Type of Each RISK FACTOR**

**Task.**

Determine the data type for each RISK FACTOR.

The data type can be continuous, binary, or categorical.

o For binary RISK FACTORS (e.g., diabetes mellitus, hypertension), specify the two possible values the RISK FACTOR can take, typically 0 (absence of the RISK FACTOR) and 1 (presence of the RISK FACTOR).

o For categorical RISK FACTORS (e.g., smoking status), specify the possible categories and their corresponding values (e.g., Non-smoker = 0, Light smoker = 1, Heavy smoker = 2).

Examples:

1. Advanced Age

o Data Type: Continuous

o Description: Age in years

2. Gender

o Data Type: Binary

o Values: 0 = Female, 1 = Male

3. Diabetes Mellitus

o Data Type: Binary

o Values: 0 = Absence of diabetes, 1 = Presence of diabetes

________________________________________

**Module 2.** Determine the Appropriate Statistical Distribution for Each RISK FACTOR

**Input:** Use the list of RISK FACTORS and their data types produced by module 1.

**Task:** Determine the Distribution Type for Each RISK FACTOR.

**Subtasks:**

**Subtask 2.1:**

o Identify the most appropriate distribution type for each RISK FACTOR, incorporating general medical knowledge specific to patients diagnosed with [DIAGNOSIS HERE] and planned to have the following SURGERY: [PLANNED SURGERY HERE].

Use the following guidelines for Choosing Distributions by Data Type:

• Continuous RISK FACTORS: Advise the selection of a distribution based on the data’s natural limits and spread within the target population:

o Normal: When values are symmetrically distributed around a mean.

o Truncated Normal: When values are concentrated near an average but are limited by realistic biological constraints (e.g., age).

o Log-normal: When values are right-skewed, common in medical data for age or time-to-event RISK FACTORS.

• Binary RISK FACTORS: Suggest using Bernoulli distributions, specifying that probability estimates should reflect the prevalence within the patient population.

• Categorical RISK FACTORS: Recommend using a multinomial distribution with proportions based on medical literature or population data if available. For example, “For smoking status, use values based on prevalence among similar patients.”

o Ensure the distribution describes the Probability Density Function (PDF) of the RISK FACTOR.

o If you do not have medical knowledge about the distribution, guess the most likely distribution.

**Subtask 2.2:**

o Select the most appropriate parameters for each distribution you identified in subtask 2.1.

o The parameters must suit the medical diagnosis of [DIAGNOSIS HERE].

o If you have no medical knowledge to support your decision, use parameters that maximize the entropy of the selected distribution.

Output format:

For each RISK FACTOR:

o RISK FACTOR NAME

o DATA TYPE

o DISTRIBUTION

o RANGE OF VALUES

o DISTRIBUTION PARAMETERS

Examples:

1. Advanced Age

o RISK FACTOR in MeSH terms: [Aged, 80 and over]

o Data Type: Continuous

o Distribution: Truncated Normal

o Range of Values: 60 to 100 years (targeting an elderly population)

o Distribution Parameters:

♣ Mean (μ) ≈ 85

♣ Standard deviation (σ) ≈ 5

♣ Lower bound = 60, Upper bound = 100

2. Gender

o RISK FACTOR in MeSH terms: [Male]

o Data Type: Binary

o Distribution: Bernoulli

o Range of Values: 0 (Female), 1 (Male)

o Distribution Parameters:

♣ Probability of Male (p) ≈ 0.6 (assuming slight male predominance in cholangitis cases)

3. Diabetes Mellitus

o RISK FACTOR in MeSH terms: [Diabetes Mellitus]

o Data Type: Binary

o Distribution: Bernoulli

o Range of Values: 0 (No Diabetes), 1 (Presence of Diabetes)

o Distribution Parameters:

♣ Probability of Presence of Diabetes (p) ≈ 0.4 (increased prevalence in elderly populations)

________________________________________

**Module 3. Generating a Cohort of Virtual Patients**

**Objective:**

- Generate a cohort of up to 50 DIFFERENT virtual patients diagnosed with [DIAGNOSIS HERE], and planned to have the SURGERY of [PLANNED SURGERY HERE].
- Each virtual patient should include values for all specified RISK FACTORS.
- **VERY IMPORTANT:** The DIAGNOSIS and PLANNED SURGERY are constants for each virtual patient. The difference between the virtual patients is in the parameters’ values of each RISK FACTOR.

**INSTRUCTIONS:**

1. Sampling

• Random Sampling:

o For each RISK FACTOR, sample a value according to its specified distribution type (e.g., Normal, Bernoulli), range, and parameters.

o Ensure sampling reflects realistic distributions coherent with the described [DIAGNOSIS], incorporating variations around the baseline clinical case.

• Clinical Plausibility:

o Check each generated virtual patient’s sampled values for clinical plausibility. For instance, ensure advanced age aligns with comorbidities like hypertension or ischemic heart disease.

o Resample any implausible combinations to maintain coherence with known profiles of patients suffering from [DIAGNOSIS HERE].

• Parameter Correlations:

o Model known correlations between RISK FACTORS (e.g., advanced age with hypertension, obesity with diabetes), especially when these correlations contribute to a higher likelihood of surgical complications.

________________________________________

2. Unique Combination Requirement

- Each virtual patient must express a **unique combination** of RISK FACTOR values within plausible ranges.
- The cohort can include **fewer than 50 patients** if fewer than 50 clinically plausible unique combinations exist.
- Ensure no duplicates across all RISK FACTOR values in the cohort.

________________________________________

**PROMPT B:** **Prediction of Complications**

**Input:**

The array of virtual patients produced by PROMPT A where each patient has a unique profile of risk factors (e.g., AcutePancreatitis, WBC, PeripancreaticInflammation).

All the virtual patients have the same:

(4) DIAGNOSIS: [DIAGNOSIS HERE]
(5) PLANNED SURGERY: [PLANNED SURGERY HERE]
(6) TIMING OF THE COMPLICATION: [TIMING HERE]

**###**

**Task Objective:**
For each virtual patient use (1) the unique profile of RISK FACTORS and (2) DIAGNOSIS, PLANNED SURGERY, and TIMING to predict the 10 most likely [TIMING HERE] surgical complications.

**Instructions for Processing the Input:**

9. Read each virtual patient’s profile.
10. Consider all available risk factors and their VALUES to identify complications **causally related to the planned surgical intervention**.
11. Use the most updated clinical knowledge for complication prediction.
12. Focus strictly on **surgical complications**—do not include general adverse outcomes or unrelated medical events.
13. For each predicted complication, identify the **most specific MeSH descriptor**.
14. Avoid returning general categories like “postoperative complications” [C23.550.767] or “intraoperative complications” [C23.550.505].
15. Ensure you predict ten complications for each of the given virtual patients.

**Output:** The output is INTERNAL and it includes the array of virtual patients, and the 10 predicted complications per virtual patient.

PROMPT C: **Summarize Complications Across Virtual Patients**

**Input:** The array of virtual patients and their complications as generated by PROMPT B. Each virtual patient contains a list of predicted complications in the following format:

[

{

"VirtualPatientID": "VP1",

"PredictedComplications": [

{"ComplicationName": "...", “NormalizedProbability": ...},

...

]

},

...

]

**##**

**Task:**

4. For each unique complication across all virtual patients, calculate:
   - VPCount: Number of virtual patients for which the complication was predicted.
   - AverageNormalizedProbability: Average of the NormalizedProbability across all virtual patients where it was predicted.
5. After calculating the averages, rescale the AverageNormalizedProbability values so that the sum across all complications equals 100.
6. Return a JSON array of dictionaries with the following fields:
   - ComplicationName
   - VPCount
   - AverageNormalizedProbability (after rescaling so the sum equals 100)

**Output format (JSON):**

[

{

"ComplicationName": "Complication A",

"VPCount": 3,

"AverageNormalizedProbability": 15.2

},

...

]

**Additional Instructions:**

- Sort the output by VPCount in descending order, then by AverageNormalizedProbability.
- Only include complications that appear in at least one virtual patient.
- Ensure that the sum of all AverageNormalizedProbability values equals 100.

\###

Output example:

[

{

"ComplicationName": "Surgical Wound Infection",

"VPCount": 10,

"AverageNormalizedProbability": 15.5

},

{

"ComplicationName": "Abdominal Abscess",

"VPCount": 10,

"AverageNormalizedProbability": 14.8

},

{

"ComplicationName": "Postoperative Hemorrhage",

"VPCount": 10,

"AverageNormalizedProbability": 12.2

},

{

"ComplicationName": "Biliary Fistula",

"VPCount": 10,

"AverageNormalizedProbability": 10.7

},

{

"ComplicationName": "Acute Pancreatitis",

"VPCount": 10,

"AverageNormalizedProbability": 10.2

},

{

"ComplicationName": "Sepsis",

"VPCount": 10,

"AverageNormalizedProbability": 8.5

},

{

"ComplicationName": "Shock, Surgical",

"VPCount": 10,

"AverageNormalizedProbability": 7.8

},

{

"ComplicationName": "Postoperative Nausea and Vomiting",

"VPCount": 10,

"AverageNormalizedProbability": 5.6

},

{

"ComplicationName": "Pain, Postoperative",

"VPCount": 10,

"AverageNormalizedProbability": 4.8

},

{

"ComplicationN
